# Human mobility-based spatial decay models of unsafe and safe water usage in rural Uganda

**DOI:** 10.1101/2025.06.27.25330423

**Authors:** Fabian Reitzug, Narcis B. Kabatereine, Betty Nabatte, Max T. Eyre, Renaud Lambiotte, Goylette F. Chami

## Abstract

Over 2 billion people lack access to safe water and are at risk of waterborne pathogens when relying on unsafe open water sites including lakes, rivers, and streams. Yet, there is limited understanding of open water usage and related human mobility for disease control. We collected 10 days of wearable GPS logger data from 452 individuals aged 5-82 years in rural Uganda. Spatial decay models were built to predict individual-level usage of open water sites and taps/boreholes to inform schistosome pathogen re-exposure and optimal tap/-borehole placement. 64.4% and 33.1% of participants visited ***≥* 1** open water site and tap/borehole, respectively. Open water site usage occurred within 10 km and tap/borehole usage within 1 km of households despite human mobility up to 100 km. Exponential decays accurately predicted site-specific open water (auROC 0.899) and tap/borehole (auROC 0.945) usage. Mobility and water site usage were non-linearly related. Open water usage was positively correlated with *Schistosoma mansoni* reinfection one year later. Even with universal tap/-borehole access, simulations showed open water usage would only decline by a maximum of 23%. Our spatial decay models offer scalable tools for identifying high-risk open water sites for pathogen transmission and optimising placement of safe water infrastructure and focal environmental interventions.

## 1 Introduction

Two key spatial processes underpin local and global transmission of pathogens: proximity to sources of infection and patterns of mobility. Proximity—typically inferred from static location information—acts as a practical proxy for exposure, capturing the likelihood of contact with infected individuals, vectors, or contaminated sites [1–3]. Mobility, in contrast, reflects dynamic movement away from the household and can reveal contacts or encounters with pathogens not captured by static data [4, 5]. While these factors are well characterised for directly transmitted infections, defining relevant contact events for environmentally mediated pathogens, especially parasites, remains challenging [6].

Epidemiological studies have often relied on aggregated mobility data to understand how pathogens spread spatially through large-scale population movements [7, 8]. This approach is particularly suited to questions about outbreak dynamics of crowd epidemic diseases such as COVID-19 or influenza, where transmission occurs via direct human-to-human contact and global spread is driven by travel [9, 10]. However, such coarse-grained representations of mobility often reduce movements to network links without explicitly accounting for spatial and environmental factors [11, 12], limiting their relevance for endemic diseases where exposure risk is highly localised. This limited relevance is apparent for tropical diseases, especially those that are environmentally-mediated where fine-scale mobility and environmental data is needed to capture how individuals interact with heterogeneous focal risk landscapes.

One example of a complex environmentally-mediated disease whose transmission is highly focal is schistosomiasis, which is caused by a parasitic flatworm and affects 250 million people globally [13]. Individuals in rural areas without access to safe water and sanitation (WASH) are most at risk; they typically acquire infection locally through contact with infested freshwater bodies such as rivers or lakes during activities such as bathing, swimming or fetching drinking water, and often remain infected for decades [14, 15]. Mathematical models have demonstrated the importance of fine-scale variation in human water contact for local transmission dynamics [16, 17] without accounting for how such fine-scale variation is shaped and constrained by the extent of human mobility. Mobility patterns of rural populations remain poorly empirically characterised [18] and existing studies on mobility as related to schistosomiasis have often relied on coarse-grained mobile phone call detail records (CDRs) [19, 20] or used no mobility data at all [21, 22]. As individual-level mobility studies using CDR data have lacked data on water contact, they have assumed proportionality between human mobility and water contact [19–21]. Consequently, it remains an open question as to whether more mobile individuals also have greater water contact and, in turn, parasite exposure. It also is unclear at which scales mobility matters for focal environmentally-dependent pathogens like schistosomes [23] and what assumptions are reasonable about mobility and water contact as a proxy indicator of exposure.

In the absence of mobility data, studies rely on household distance to open water sites as a proxy under the assumption that people visit the closest water site to their home [24, 25]. Yet, personal preferences, site characteristics or social norms may lead individuals to travel further [26].

Human mobility is important for schistosomiasis control efforts such as environmental control (mollusciciding) and WASH, recommended by the 2022 WHO guideline [27]. It is assumed that safe water supplies such as taps/boreholes reduce exposure [27]. However, without detailed data on water usage, it remains unclear how much open water contact may be diverted and how far people travel to use safe water infrastructure. Here, water usage is defined here as i) usage of open water sites (lakes, rivers, streams, ponds) and ii) usage of taps/boreholes. The former is considered unsafe and the latter safe within the context of schistosomiasis [14]. For mollusciciding, the two main strategies are focal control (treating all open water sites used by the community to kill freshwater snails, the intermediate host of the *Schistosoma* parasite) and radius control treating all sites within 500 metres (m) of human settlements [28]. There are no available mathematical or empirical models to predict water site usage patterns over distance to aid in identifying the optimal mollusciciding strategy.

We conducted a large-scale community-based study using wearable GPS loggers to combine individual-level movement data with detailed field mapping of open water sites and safe water infrastructure. 452 individuals were sampled within the SchistoTrack Cohort in rural Uganda [29]. After treatment with praziquantel, individuals were tracked for ten days between January-February 2022 with GPS points recorded at two-minute intervals between 5 a.m. and 8 p.m. *S. mansoni* infection status and intensity before treatment, and reinfection intensity one year later were measured and correlated against water usage. To establish a proxy indicator of human schistosome exposure [24], we focused on human contact with shallow water sites (<3.7 metres [m] in maximum depth), henceforth called open water sites, where parasite density due to snails of the species *Biomphalaria sudanica* and *B. stanleyi* has been shown to be greatest [30]. We statistically assessed the roles of distance and mobility as predictors of human water contact by developing a series of spatial decay models, which are individual-level formulations of the gravity model [31, 32]. The primary aim of our study was to build individual-level spatial decay models that use human mobility data to predict fine-scale water usage patterns. Our secondary aim was to provide simulations that quantify the extent to which open water site usage and frequency may be reduced due to the usage and availability of safe water infrastructure.

## 2 Results

### 2.1 Measures of open water contact and human mobility

We derived indicators of open water usage and taps/borehole usage as shown in Fig. 1. Within our study catchment, we separately clustered the 143 open water sites and 63 taps/boreholes mapped by SchistoTrack into 69 distinct open water sites and 32 tap/- borehole locations. A human contact event with an open water site or a tap/borehole was defined as an individual moving within a 30m buffer of the location of interest, comparable to buffer sizes (20–30m) used in other GPS logger studies [33, 34].

**Fig. 1:**
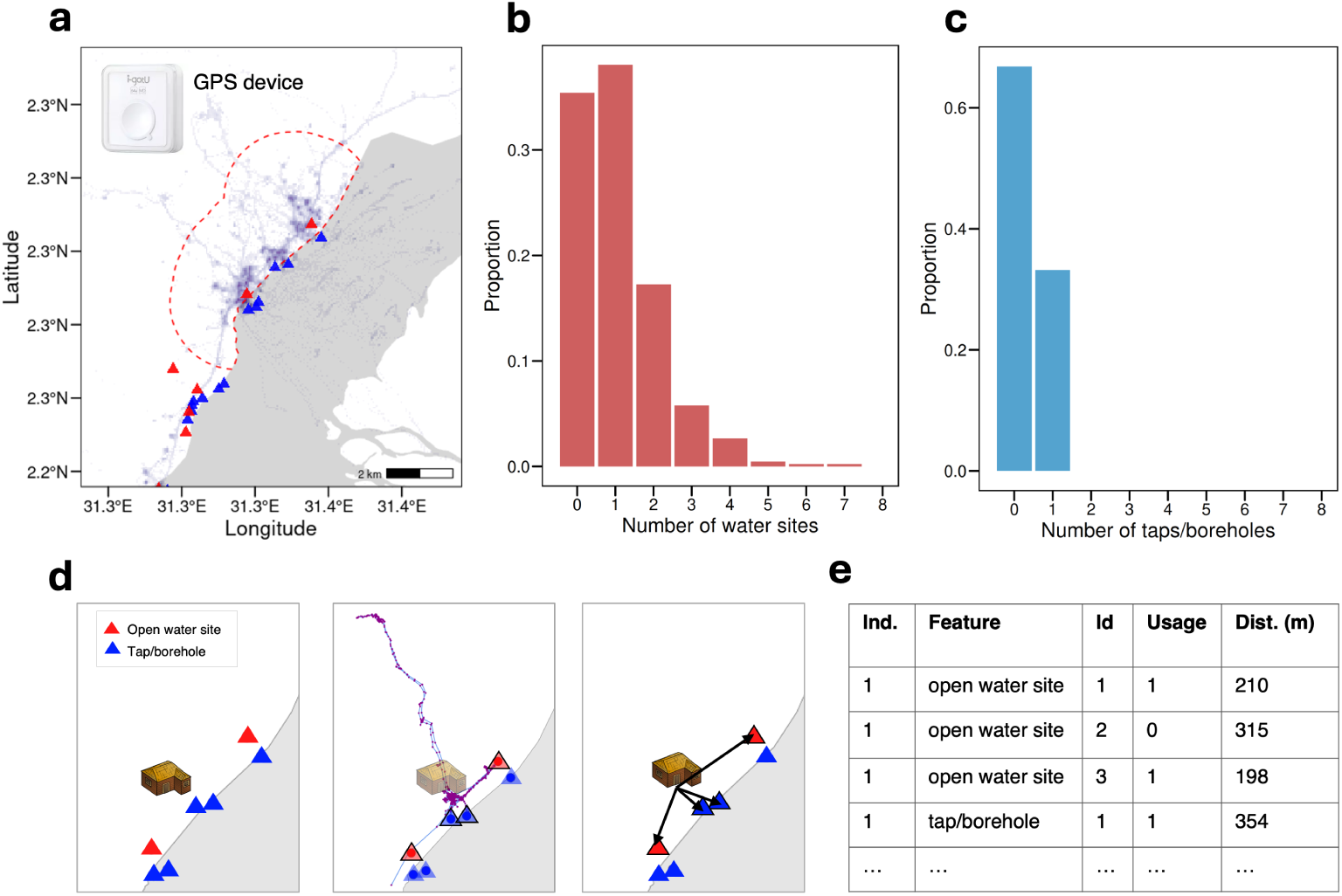
GPS logger data and processing. **a** GPS logger data traces in Pakwach aggregated across the study sample (GPS locations/movement represented here as purple lines). Blue triangles denote open water sites mapped by the study team. Red triangles denote taps/boreholes. The red dashed line shows the study area. **b** Number of distinct open water sites visited by study participants. **c** Number of distinct taps/- boreholes visited by study participants. **d** Processing steps to derive open water site and tap/borehole usage data from GPS data and household and site location data. **e** Data frame resulting from the processing which is used to train spatial decay models.

As we were interested in the movement of an individual relative to the home location for visits to open water sites, taps/boreholes, and any other purpose (’general mobility’), we focused on the radius of gyration R*_g,i_* (equation 1) to capture mobility as this provides a measure of displacement of an individual i from the household location and is widely used in human mobility modelling [32, 35, 36].

### 2.2 Patterns of open water contact and tap/borehole usage

There were 25 distinct water sites in Pakwach, 26 in Buliisa, and 18 in Mayuge with a total of 8249 water contact events observed across districts during the study. 64.4% (292/452) of all participants visited at least one water site. The median duration of water contact for all participants was 3 minutes (min) per day (IQR: 0–13.3 min). The distribution of different sites visited per individual is shown in Fig. 1. Among the 292 individuals who had water contact, the median number of different water sites visited was 1 (IQR: 1–2) and the median daily duration was 9 min (IQR: 3–23 min). The number of participants using at least one water site varied across districts (χ-squared statistic 76.8, df = 2, p < 0.01) with 53.6% (98/183) in Pakwach, 90.2% (148/164) in Buliisa, and 46% (46/105) in Mayuge. There were differences in water contact patterns throughout the day by district. In Pakwach, open water contact peaked at 8:00 a.m., whereas in Buliisa and Mayuge, it peaked around 7:00 p.m. (Fig. S1a).

There were two boreholes and five taps in the study villages in Pakwach; three boreholes and four taps in Buliisa; and two boreholes and 16 taps in Mayuge. The price for using these facilities was the same across districts; borehole usage cost UGX 1000 (USD ∼ $0.27) per household per month and taps cost UGX 100 (USD ∼ $0.03) per jerrycan. A total of 1181 tap/borehole usage events were observed, which was 85.7% fewer than the number of water contact events. Among all study participants, 33.1% (150/452) visited at least one tap/borehole. The median duration of tap/borehole usage across all participants was 0 min per day (IQR: 0–1 min). The distribution of distinct taps/boreholes visited is shown in Fig. 1. Among the 152 individuals who used taps/boreholes, the median daily duration spent retrieving water was 3 min (IQR 1– 10 min), and the median number of different taps/boreholes visited was 1 (IQR: 1–1). Usage was highest in Buliisa, where 49.4% (81/164) of participants used at least one tap/borehole, followed by 25.1% (46/183) in Pakwach and 21.9% (23/105) in Mayuge. Across all districts, tap/borehole usage peaked between 5:00–7:00 p.m. (Fig. S1b).

Overall, 24.8% (112/452) of participants visited both water sites and taps/boreholes and 27% (122/452) visited neither. Usage of water sites and taps/boreholes was weakly positively correlated (Spearman rank correlation [ρ*_s_*] = 0.15, p < 0.01). There also was a weak positive correlation between the duration of water site and tap/borehole usage, measured in min per person per day (ρ*_s_* = 0.17, p < 0.01). However, among only participants (330/452) with either water site or tap/borehole usage, there was a negative correlation between the duration of water site and tap/borehole usage (ρ*_s_* = −0.12, p = 0.02).

### 2.3 Relationships of age, gender, and household distance with water usage

Study participants were aged 5-82 and 50%(226/452) were female. In terms of demographics, we found no significant effects of age and gender on open water site or tap/borehole usage (Table S1). Living in a household with an improved drinking water source such as taps/boreholes was positively associated with both open water site and tap/borehole usage. Occupation was relevant to open water site usage as fishermen and fishmongers were more likely to use open water sites (Table S1). Consistent with self-reported measures of human water contact in the larger SchistoTrack study [24], open water contact peaked around age 25, whilst tap/borehole usage was largely stable across age (Fig. S2). Water usage was correlated between adults and children within the same household (ρ*_s_* open water sites = 0.34, p < 0.01, n = 261, ρ*_s_* taps/boreholes = 0.34, p < 0.01, n = 261).

As SchistoTrack focused on fishing villages, households lived close to waterbodies with a median distance of 225m (IQR: 82–412m) to the closest open water site. The median distance to the closest tap/borehole was 251m (IQR: 93–664m). Fishermen lived closer to open water sites (median 84m, IQR 58–190) compared to other adults (median 233m, IQR 74–400, Wilcoxon rank-sum test statistic [*W* ] 2599, p < 0.01). Households residing closer to open water sites were more likely to be closer to taps/boreholes as well (ρ*_s_* = 0.52, p < 0.01, n = 261).

Across all study participants, individuals travelled slightly further to visit open water sites compared to taps/boreholes, though the difference was not statistically significant. The median distance to a frequented open water site was 229m (IQR 107– 420m), whereas the median distance to a frequented tap/borehole was 172m (IQR 123–320m, Fig. S2c–d; *W* 38566, p = 0.15). For open water contact, the closest site to the home of an individual accounted for 47% (224/477) of all distinct sites used by individuals, and for 61.9% of total observed usage duration (58152/93865 min). The closest tap/borehole to the home of an individual accounted for 99.3% (149/150) of all distinct taps/boreholes used by individuals and for 99.6% of the total usage duration (16417/16447 min).

### 2.4 Relationship of human mobility with water usage

Aggregate human mobility patterns by district are shown in Fig. 2a. These patterns were well approximated by a truncated power-law distribution (r^2^ = 0.73) within the range of ∼ 1.3 × 10^1^ − 10^4^ m. A district-specific model performed better than a global model (likelihood ratio test p < 0.01). Overall, the median radius of gyration among participants was 380m (IQR 151–1157m). Although not significantly different due to the high variability of movement within districts, participants in Pakwach had the greatest area of movement with a median radius of 633m (IQR 228–1454m), followed by individuals in Mayuge (median 304m, IQR 152–698m) and Buliisa (median 262m, IQR 93–1222m). Fishermen (median 658m, IQR 187–1757m) were not substantially more mobile than other adults (median 506m, IQR 134–1478m). Due to our sampling design being stratified by district (Fig. 2), we grouped participants into district-specific terciles of mobility (bottom, middle, top; see Fig. 2b) for subsequent analyses. When comparing the upper limits of general mobility and visits to water sites and taps/boreholes, tap/borehole visits were more spatially focal than water site visits and general human mobility. Virtually all tap/borehole visits were within 1000m, whereas all water site visits were within 10 km, and, at the more extreme end, all human movement was within 100 km from the household (Fig. 2c). Individuals who lived >1000m from water sites had a greater median radii of gyration (median 620m, IQR 311–1260m) than those living within 0–500m (median 349m, IQR 143–1083m, Fig. 2d), though not statistically significant due to the high level of individual variation.

**Fig. 2:**
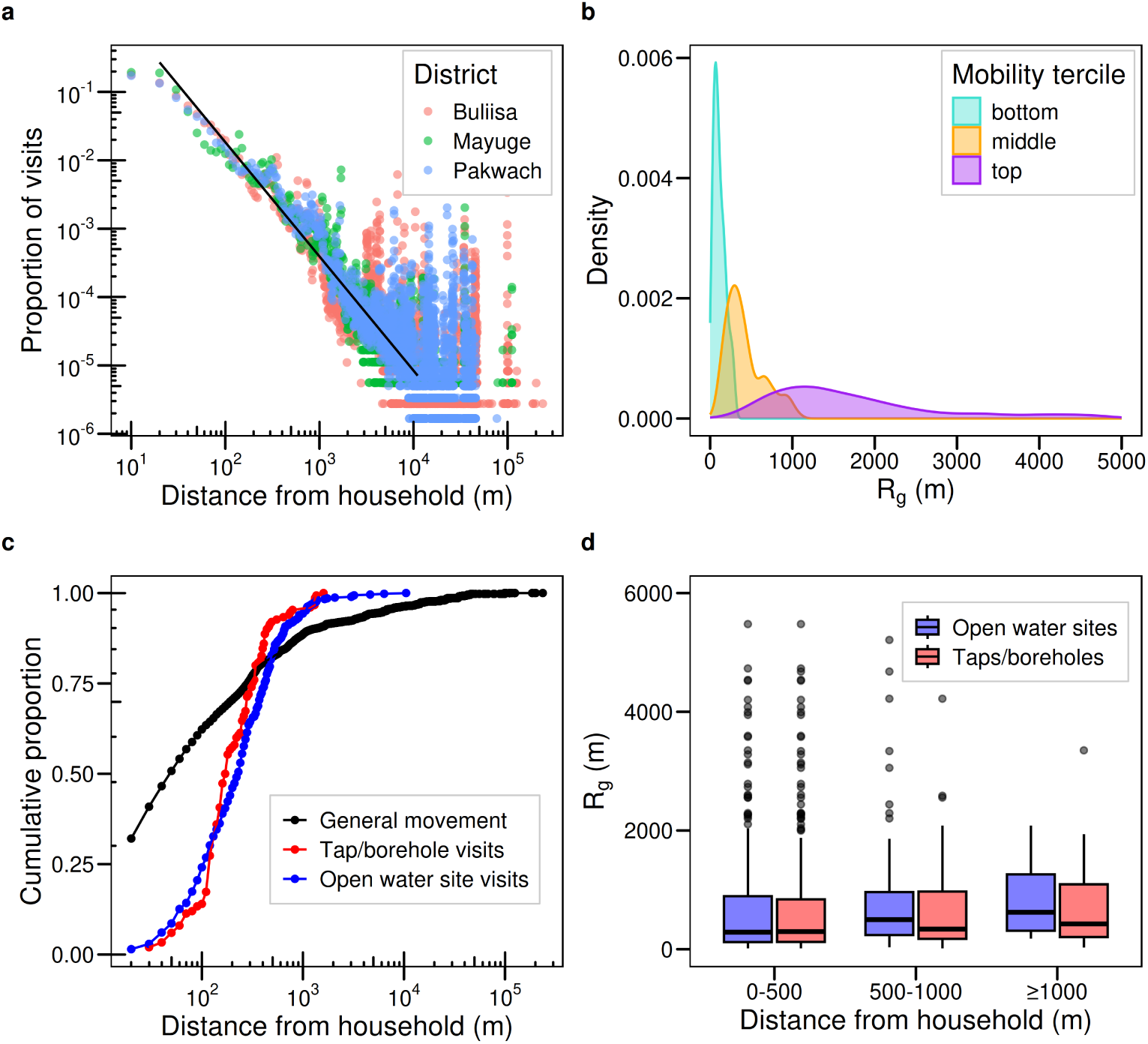
The relationship between mobility and usage of open water sites and taps/boreholes. **a** Proportion of visited locations over distance from the household. The black line represents a truncated power law distribution fitted between ∼ 1.3 × 10^1^ − 10^4^ m. Limits were determined using the poweRlaw R package. **b** Distribution of radii of gyration (R*_g_*) (eq. 1) by district-specific mobility tercile. R*_g_* measures human mobility relative to distance from the household. **c** Cumulative proportions of visits to open water sites, tap or borehole locations, and all GPS locations (’general movement’), plotted against distance from the household **d** Relationship between R*_g_* and household distance to the nearest open water site and tap/borehole.

### 2.5 Spatial decay models of water usage

Fig. 3 introduces four individual-level spatial decay models built to predict which water sites individuals were likely to use focusing on household distance only (Fig. 3a), household distance plus choice of water sites, (Fig. 3b), household distance and human mobility (Fig. 3c), and household distance and the presence of taps/boreholes, which may crowd out open water site usage (Fig. 3d). Spatial decay models were fitted as Bayesian non-linear regression models to predict the probability of an individual using a specific open water site and tap/borehole (see equation (eq.) 2 and Methods for details). Models were trained on 85% of the full data and evaluated by calculating the area under the receiver operating curve (auROC) on 15% held-out data.

**Fig. 3:**
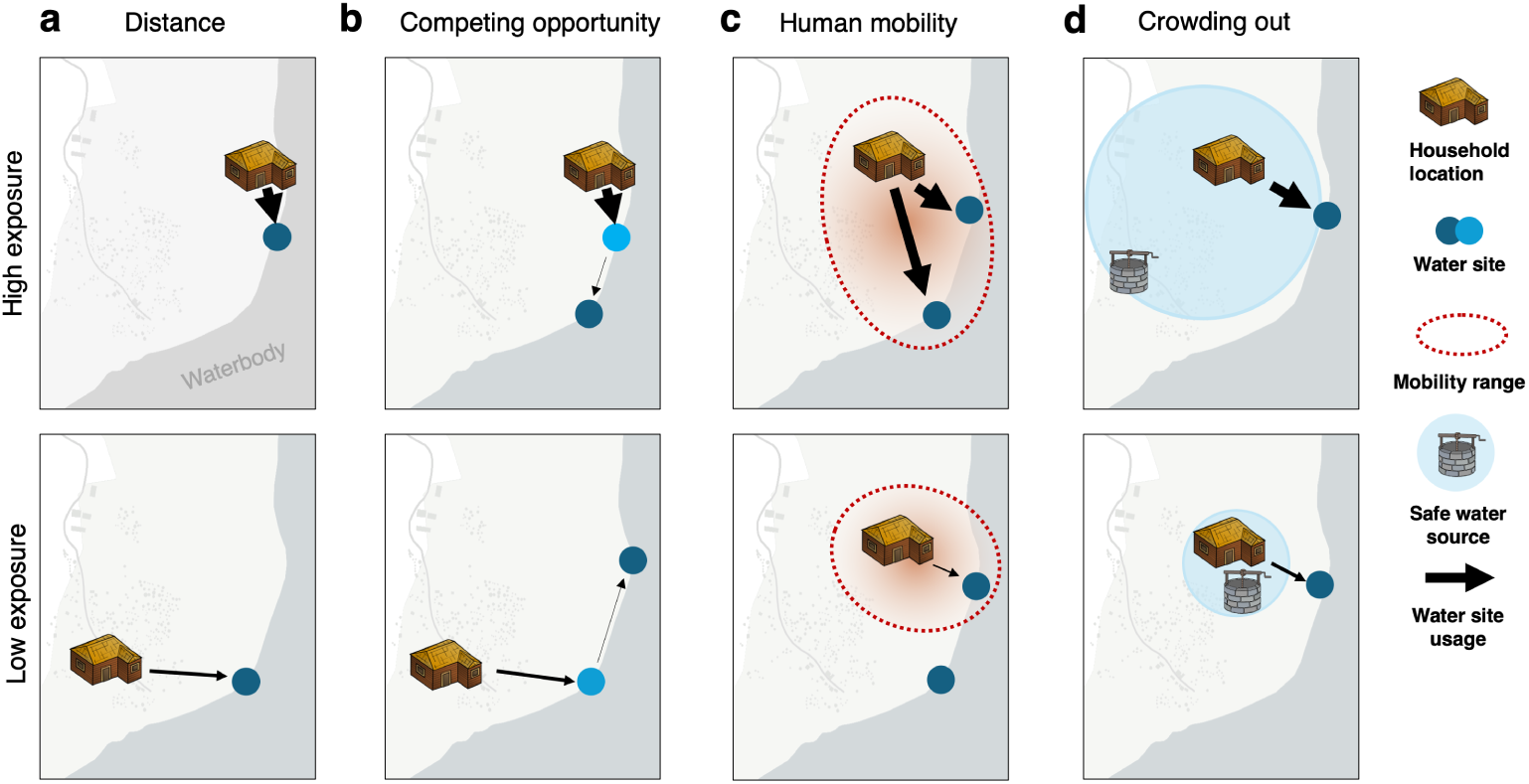
Schematic of different spatial factors influencing open water site usage. **a** Open water site usage depends purely on household distance to the site. **b** Open water site usage depends on household distance to the index site (dark blue) plus distance to competing sites (light blue). **c** Open water site usage depends on the range of human mobility. **d** Open water site usage depends on proximity to safe water infrastructure such as boreholes, whereby boreholes are able to reduce (i.e. crowd out) open water contact.

An exponential decay best approximated open water site usage (Fig. S3). The exponential spatial decay model performed better than a simple logistic regression model using household distance, especially for predicting open water site usage at small spatial scales (< 100m, Fig. S4). Distance-based models outperformed all other logistic regression models using socio-demographic and behavioural proxy variables to predict open water site usage, based on auROC (Table S2).

The global spatial decay model of open water site usage is shown in Fig. 4a. This model had an auROC of 0.877. For individuals living adjacent to open water sites (20m), the probability of open water site usage was 70% (95% credible interval [CrI]: 61–79%). The probability of open water site usage reduced to 51% (95% CrI: 46–57%) at 100m, 11% (95% CrI: 10–12%) at 500m, and 1% (95% CrI: 0.7–1.4%) at 1127m (Fig. 4a).

**Fig. 4:**
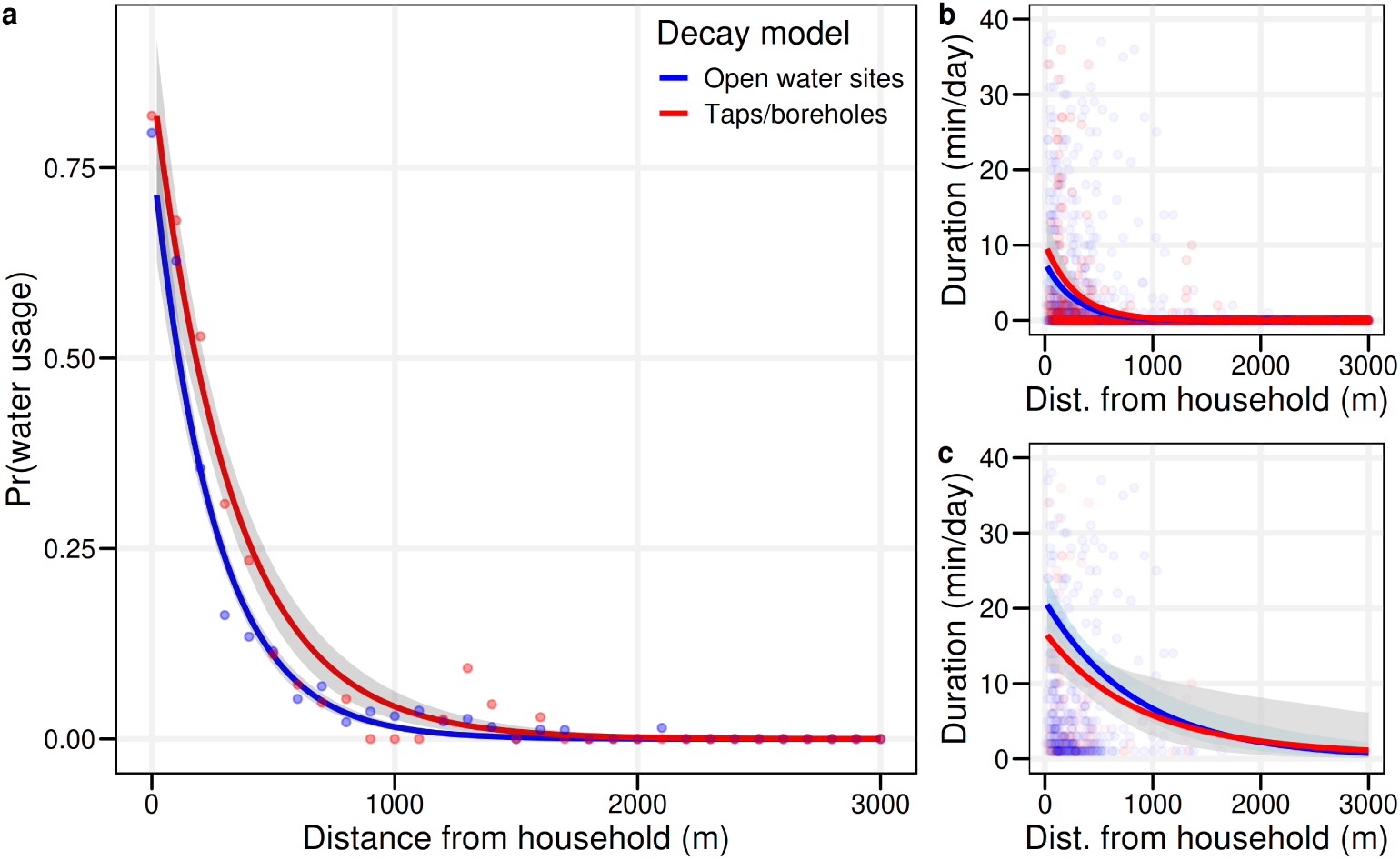
Spatial decay baseline models. **a** Individual-level spatial decay models predicting the probability of water site and tap/borehole usage over household distance. Dots represent the data. **b** Spatial decay models predicting the duration of water site and tap/borehole usage over household distance among all individuals. **c** Spatial decay models predicting the duration of water site and tap/borehole usage over household distance, restricted to individuals using water sites or taps/boreholes, respectively. Shaded grey areas represent 95% credible intervals.

The spatial decay for tap/borehole usage was similar to the decay observed for open water sites (Fig. 4a). The spatial decay model for taps/boreholes had an auROC of 0.932. For individuals adjacent to taps/boreholes, the estimated probability of any usage was 83% (95% CrI: 70–92%), 12%-points higher than for open water sites. The estimated probability of use was 64% (95% CrI 56–71%) at 100m, 18% (95% CrI: 15– 22%) at 500m, and 1% (95% CrI: 0.5–1.9%) at 1448m, compared to 1127m for contact with open water sites (Fig. 4a). For both open water site and tap/borehole usage, we tested the inclusion of additional covariates and found that district-specific decay models performed best (auROCs of 0.888 and 0.945). No other covariates, including age category, occupation, and self-reported safe/unsafe water contact behaviours, substantially improved model performance (Table S3).

We also used exponential spatial decay models to predict the duration of open water site and tap/borehole usage (Fig. 4b-c). The duration models had moderate predictive performance (ρ*_s_* open water sites = 0.36 and ρ*_s_* taps/boreholes = 0.47). Overall, open water site usage duration and probability of any usage had similar decays (both decay parameters [b_1_] 0.004). Among users, the decay in open water site usage duration was less pronounced than across all participants, as predicted durations remained positive even at 2000m (2.2 min [95% CrI 1.0–4.4], Fig. 4c).

### 2.6 Competing opportunities and the role of human mobility

All previous models assumed that the influence of distance on water usage was not influenced by the number of opportunities available. However, if two individuals reside 40m from an open water site, their contact patterns may be different if one individual has access to a site at 20m already. Incorporating such competing opportunities improved the predictive performance of the spatial decay models of open water site usage (hypothesis b in Fig. 3). Exponential spatial decay models with competing opportunities are shown in Figs. 5a-b. There was a monotonic decay in the probability of open water site usage (Fig. 5a) by the sequence of sites, ordered by their relative distance from the home of an individual. This model outperformed the best-performing model from the previous section, a district-specific decay model (auROCs of 0.899 and 0.888, respectively). A simpler competing opportunity model which relied only on site order without regard to the household distance performed less well (auROC of 0.845, Fig. S5).

**Fig. 5:**
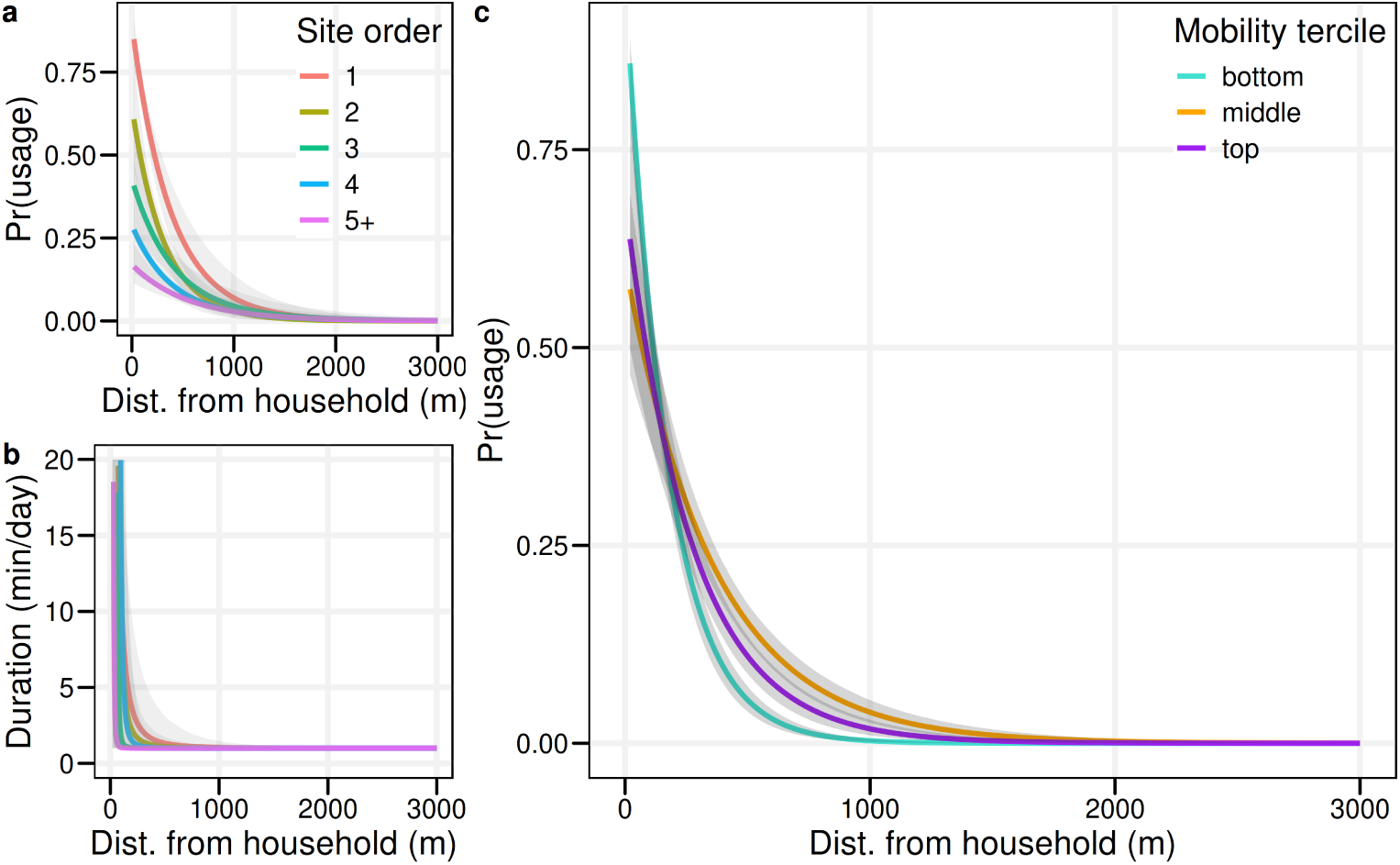
Spatial decay models incorporating human mobility and competing opportunities. **a** Spatial decay model predicting open water site usage probability based on site order. For each individual, we ranked the sites by their proximity to their home, identifying the closest site, second closest, third closest, and so on, up to the fifth furthest site. (All open water sites beyond the fifth site were grouped into the fifth closest site category). Separate decay models were then fitted for each of these site ranking categories. **b** Spatial decay model predicting open water site duration based on site order. **c** Spatial decay model predicting open water site usage probability based on district-specific mobility tercile. Shaded grey areas represent 95% credible intervals.

We also investigated whether individuals with higher relative levels of mobility tended to visit open water sites at greater distances (Fig. 3c). There was a nonmonotonic effect of mobility on open water site usage whereby individuals in the middle district-specific mobility tercile maintained the highest site usage probabilities across distance. Surprisingly, both individuals in the high and low mobility terciles had similar, more pronounced decays over distance (Fig. 5c). The decay model incorporating human mobility also outperformed the district-specific decay model (auROCs of 0.893 and 0.888, respectively). We repeated this analysis for taps/boreholes and found that incorporating mobility did not improve model performance, compared to the district-specific decay model for taps/boreholes (auROCs of 0.909 vs. 0.945, respectively).

### 2.7 Reductions in open water contact due to safe water infrastructure

To investigate whether crowding-out effects exist where open water site usage is fully or partially substituted by tap/borehole usage (hypothesis d in Fig. 3), we built spatial decay models in which usage of taps/boreholes reduces the probability and frequency of open water site usage (see Methods).

We found that the crowding-out model (hypothesis d in Fig. 3) slightly outperformed the model without crowding out (auROCs of 0.890 and 0.888, p = 0.02). The crowding-out parameter α had a value of 0.30 (95% CrI: 0.04–0.56), suggesting that taps/boreholes might reduce the probability of open water site usage by 30%. The overall effect on open water site usage was small because a 30% reduction was only achieved for individuals with open water contact who also lived directly next to taps/boreholes (20m). The crowding-out model suggested that in our sample < 2% of individuals (5/452) were predicted to have no open water contact due to the presence of taps/boreholes (95% CI: −15–74 individuals) based on comparing observed open water contact patterns to a counterfactual scenario without any taps/boreholes. Reductions in open water contact were unevenly distributed across the population. Individuals with the highest predicted reductions in open water site usage probabilities tended to have the highest tap/borehole usage probabilities, which resulted in the largest crowding-out effects (Fig. 6d).

**Fig. 6:**
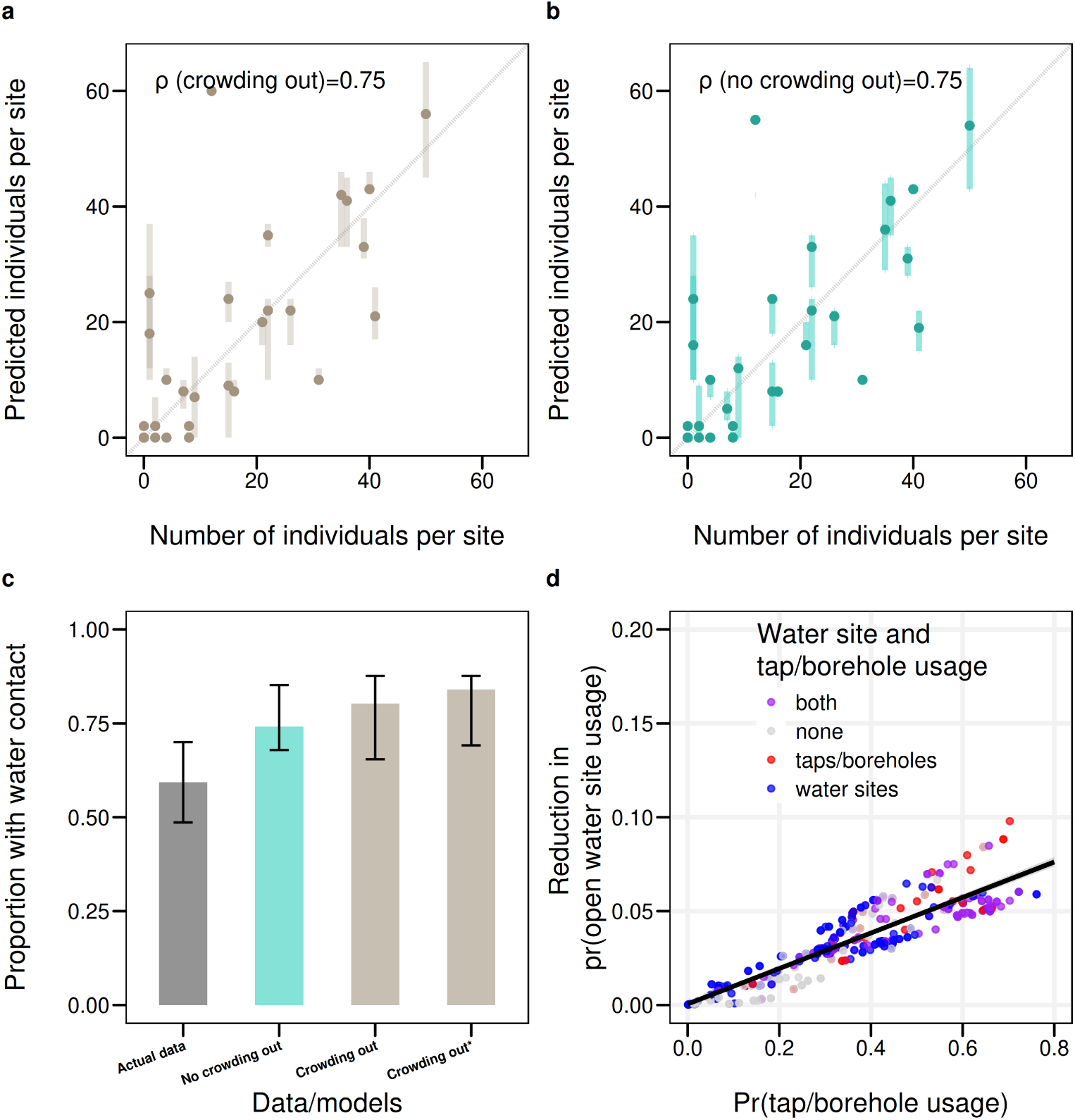
The influence of taps/boreholes on open water site usage probability. **a-b** Predicted number of individuals using each water site versus the true number. **c** True proportion of individuals with water contact versus the proportion predicted in spatial decay models with and without crowding out. The ‘crowding out^*^ model’ represents a model in which the proportion of individuals was estimated under a counterfactual scenario where no taps/boreholes were present. Thus, the difference between the ‘crowding out’ versus ‘crowding out^*^’ models represents the estimated impact of taps/boreholes on water site usage. **d** Scatter plot showing the predicted probability of tap/borehole usage versus the predicted probability of water site usage for each individual. The length of the arrows indicates the reductions associated with the presence of taps/boreholes for each individual. Colours represent actual usage of water sites and taps/boreholes for each individual. Again, this was estimated by comparing the observed values for tap/borehole coverage to a counterfactual scenario where no taps/boreholes were present. Vertical bars in **(a–c)** represent 95% confidence intervals. The colours in **(d)** represent the true usage patterns for each individual.

We repeated the crowding-out analysis for duration of open water site usage (Fig. 7). We found again that the crowding-out model performed slightly better in predicting the observed durations of water contact compared to a model without crowding out (ρ*_s_* = 0.38 versus ρ*_s_* = 0.37). For duration, the model suggested that existing taps/boreholes had a crowding-out effect of 33% (95% CrI: 0.02–0.74), which was slightly larger than the effect on the probability of any open water contact (30%). The average duration reduction was 0.5 min or 34% (duration of 1.36 min (95% CrI: 0.67–3.34) without taps/boreholes and 0.9 min (92% CrI: 0.57–1.52) with taps/boreholes. Almost all reductions (>95%) were concentrated among the individuals in the top three deciles of duration of open water site usage (Fig. 7b). Even under universal tap/borehole usage, open water site usage duration could only be reduced by additional 23% (95% CrI: 19–37%) (Fig. 7c).

**Fig. 7:**
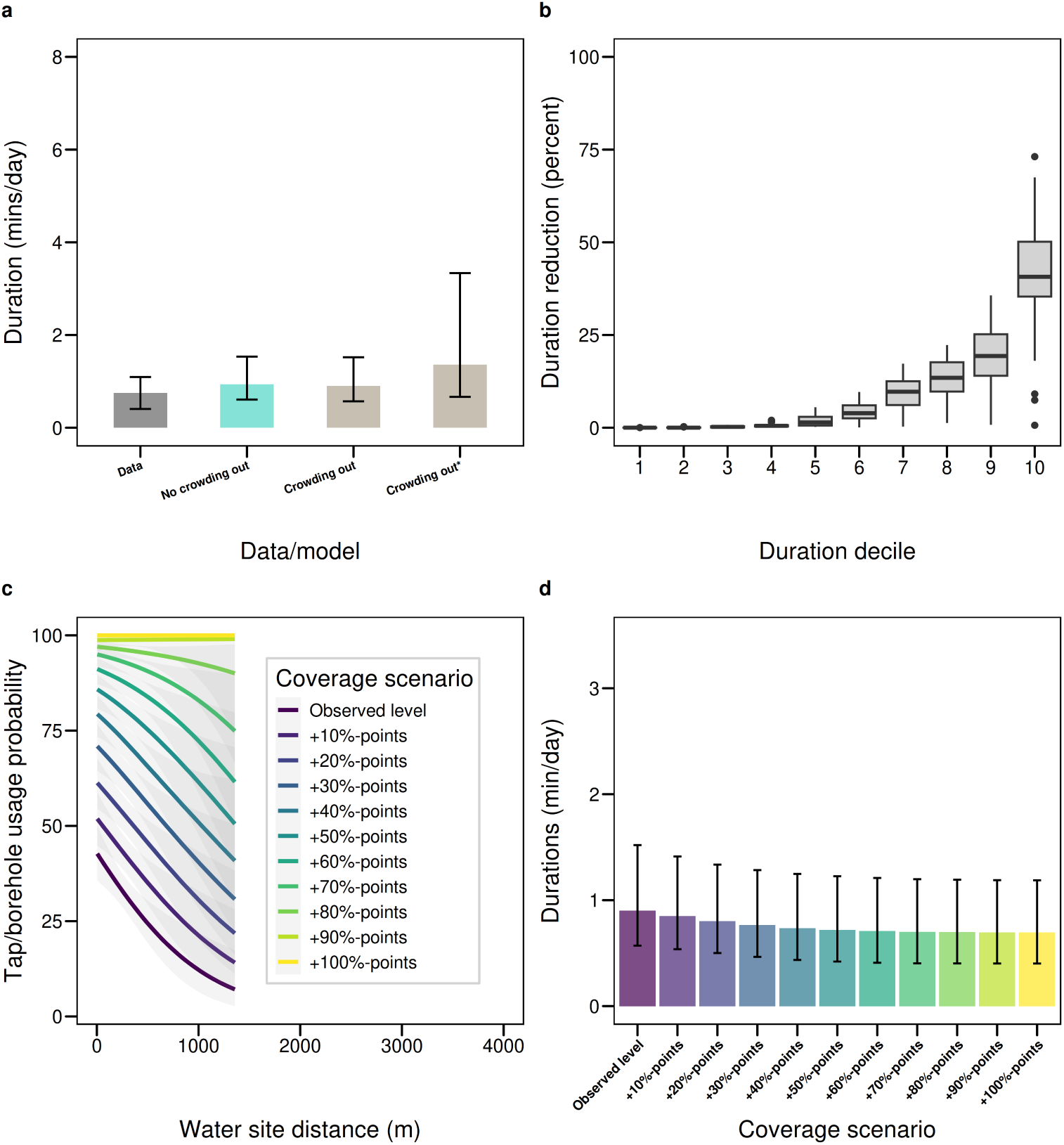
The influence of taps/boreholes on open water site usage duration. **a** True duration of open water contact per site in min per day versus estimated durations predicted using the crowding-out and no crowding-out models. The ‘crowding-out^*^ model’ represents a model in which open water site usage duration was estimated under a counterfactual scenario where no taps/boreholes were present. **b** Relative reductions in water site usage duration by duration decile. **c** Tap/borehole usage probability estimated from spatial decay model, (black line) and ten different hypothetical scenarios where the tap/borehole usage of individuals was increased by 10–100%-points from the observed level (with 100%-points representing universal usage of taps/boreholes). **d** Estimated relative reductions in open water site usage duration under the scenarios shown in **(c)**. Vertical bars in **(d)** represent 95% credible intervals.

As people with the highest durations of open water contact tended to have high levels of tap/borehole usage (Fig. 7c), the potential to further crowd out their open water contact with safe water supplies was limited. Nonetheless, crowding out potential was highest near water sites, as we estimated that for households, within 100m of water sites, 46% of open water contact duration (95%CI: 25–64%) could be crowded out, whilst this number was 7% (95%CI: 3–14%) at 500m and 1% at (95%CI: 0.5–3%) at 1000m (Fig. S6).

### 2.8 Applying spatial decay models to suggest locations for environmental control and WASH interventions

Given that our model was highly predictive of the true number of people visiting each water site (ρ = 0.75, Fig. 6a-b), we applied this model for out-of-study predictions, focusing on Pakwach by extracting footprints of all 5177 local structures in the catchment around the study villages under the assumption that each building represented one household (see Methods). The results in Fig. 8a suggested that there were substantial differences in open water site usage even across nearby sites with one site predicted to be used by 11 households and another by 96 households.

**Fig. 8:**
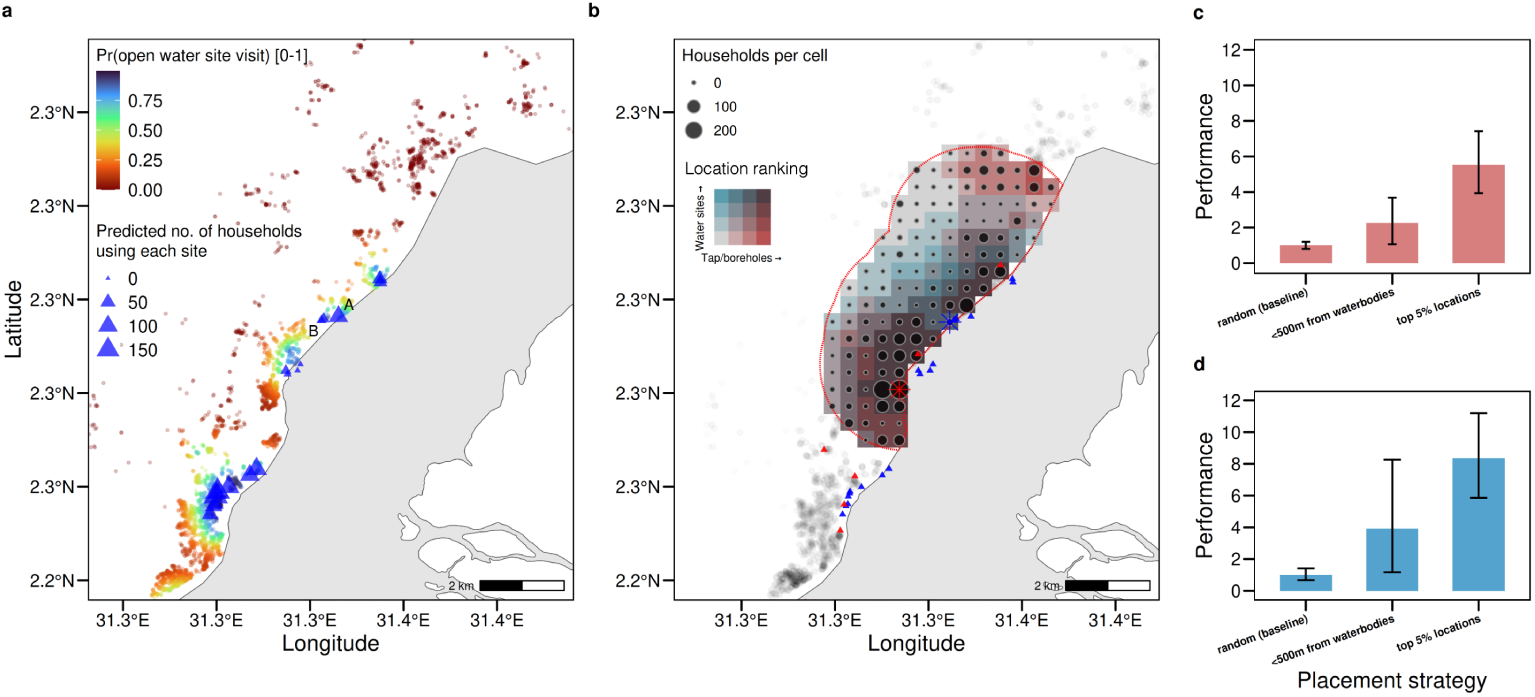
Applying spatial decay models to predict population-level water site usage and optimal locations for safe water infrastructure. **a** Populationwide predictions of the water site usage probability for each household based on 5177 buildings in Pakwach (each dot represents one building). Colours of the dots represent the predicted probability of using one or more open water sites. Blue triangles show all local water sites mapped by the SchistoTrack study, and their respective sizes indicate the number of households predicted to use each site (multiple assignments possible). Labels ‘A’ and ‘B’ denote two nearby open water site predicted to have substantially different numbers of households using each (96 and 11, respectively). **b** Optimal location for building a new borehole, estimated using simulations on a 400m × 400m grid for all locations within the dashed area (within 1000m of study households). All locations were ranked for each of the two strategies. Darker colours represent higher ranked locations for placing taps/boreholes (dark red, dark blue, and brown represent locations ranked highly based on strategy i, strategy ii, and both, see text for details). The red and blue star symbols represent the optimal locations for strategy i and strategy ii, respectively. **c–d** Performance of the top 5% of locations for two placement strategies compared to random placement, or placement within <500m of waterbodies. Vertical bars in **(c–d)** represent bootstrapped 95% confidence intervals.

To suggest the optimal location for building a public borehole, we evaluated two different tap/borehole placement strategies using simulations, maximising either population-level safe water coverage (strategy i) or crowding out of open water contact (strategy ii, Fig. 8b). Model-based placement of taps/boreholes was estimated to substantially improve the benefits of safe water infrastructure. We estimated that population-level usage probability was 5.5 times as high in the top 5% of locations compared to random placement. Similarly, population-level crowding out achieved at the top 5% of locations was 8.3 times as high compared to random placement (Fig. 8c-d). Maximising population-level coverage versus crowding out suggested slightly different optimal locations (star symbols in (Fig. 8)b).

### 2.9 Model validation against reinfection data and sensitivity to removing short contact events

We assessed whether our model results were relevant for schistosome reinfection by correlating *S. mansoni* reinfection intensity with predicted and actual open water site usage duration. The correlations between our model-based estimates of duration (eq. 4) and actual durations with reinfection were similar (ρ*_s_* = 0.21 [0.95% CI 0.13–0.30] and ρ*_s_* = 0.23 [0.95% CI 0.15–0.31]). We assumed our model approximated general water site usage patterns historically and at-present, and that human water contact is relevant for acquiring new infections.

For minimal data needs and GPS recording frequency, only 25 individuals recorded over ten days or 452 individuals recorded over one day were sufficient to train the decay model (Fig. S7a). Concerning spatial generalisability, training data on two districts and predicting the decay for the held-out district achieved auROCs>0.82 when holding out each district. Our decay parameters b_0_ and b_1_ were robust to removing all open water contact events of <5 min or <10 min duration, respectively (Fig. S8). Additional robustness analyses (Appendix A.1) supported our results including describing any differences between participants with complete versus incomplete GPS logger data (Table S4), investigating wear non-compliance (Table S5), and assessing systematic biases in predictions of spatial decay models across socio-demographic characteristics (Table S6).

## 3 Discussion

The WHO schistosomiasis guideline emphasises the need for WASH, environmental control and behaviour change interventions to reduce schistosome exposure in endemic areas [27]. However, due to the reliance on self-reported water contact data or coarse mobility indicators [15, 19], our understanding of human behaviour relevant for exposure remains insufficient for intervention design. Here, we used wearable GPS logger data to predict fine-scale water usage patterns of 452 individuals in rural Uganda. We developed a novel suite of individual-level spatial decay models for open water site usage and tap/borehole usage, identifying exact sites used by individuals across a wide age range of 5-82 years. We demonstrated the utility of spatial decay models for finescale prediction of local human exposure, validated for their relevance to schistosome reinfection.

Our individual-level models performed well in rural Uganda at predicting open water site and tap/borehole usage (auROCs of 0.899 and 0.945, respectively). This finding is in contrast to past studies that used aggregate mobility data and found that gravity models performed poorly in rural areas [37, 38]. Previously, studies used only coarse mobility data from cell phones [19, 20], no mobility data at all [21, 22], or relied on small GPS logger samples [33, 39] to predict general water contact. Our models demonstrate how simple exponential spatial decay models can approximate small-scale environmental exposures. A similar decay pattern has been suggested for water usage within the context of trachoma [40].

We found no clear socio-demographic correlates of human mobility. There has been increased interest in identifying mobile individuals who may be missed in mass drug administration (MDA) programmes [41, 42]. In our study, human mobility did not substantially vary based on gender or household proximity to waterbodies. Adults were found to have a substantially larger range of movement compared to children, though the median mobility was not significantly different. Fishermen, who have been thought to be especially mobile [43], did not have a larger range of movement than other adults. As most fishermen lived close to the shoreline, they had to travel shorter distances to access open water sites compared to the general population. These results suggest a need for a formal analysis or mathematical model of how mobility actually may impact MDA coverage or transmission reduction efforts, given that past assumptions of mobility around high-risk individuals (fishermen) were not upheld.

Contrary to what has been assumed elsewhere, open water contact was not proportional to human mobility, as captured by the radius of gyration [20, 21]. In our study, people with intermediate levels of mobility travelled further to access water sites, compared to people in the highest and lowest tercile of mobility. Open water contact of low mobility individuals, for instance children, might have been constrained by limited movement, while highly mobile individuals may have moved far for reasons unrelated to open water usage. Most importantly, the spatial scales of water usage and mobility differed. People travelled up to 1 km to visit taps/boreholes, up to 10 km to visit open water sites, and up to 100 km for any purpose with different spatial decays for human mobility versus open or safe water use. While general mobility was approximated by a truncated power law, consistent with other human mobility datasets [35, 36], water usage (open water or tap/borehole) was best modelled using an exponential spatial decay, highlighting that spatial patterns of mobility cannot serve as a direct proxy of water contact. Thus far, mathematical transmission models are not spatially explicit [44, 45]. Our results provide empirical data which could be used to parametrise such models with human movement and water contact information.

Our simulations suggest, at best, moderate reductions of open water contact due to improved safe water provision with no level of safe water provision that would eliminate all open water contact. The most optimistic estimate suggested that existing taps/boreholes reduced water contact duration by 33% compared to a hypothetical scenario where no taps/boreholes were present. As individuals in this study lived close to taps/boreholes (median of 251m), we found that even under universal safe water usage, the ability to further reduce their open water contact duration was capped at 23% (with daily levels already very low, <4 min). We estimated that the presence of taps/boreholes completely eliminated open water contact for only <2% of individuals. However, even if effects of safe water on overall open water contact levels—the focus of this analysis—are low or non-existent, it remains possible that specific activities, such as getting drinking water, still can be addressed [24], especially given that most diverted open water contact was found for individuals already with a high probability of tap/borehole usage. There exist conflicting findings as to what extent safe water access can reduce schistosome infection [14, 27, 46]. Our results suggest caution in assuming that the provision of safe water would reduce schistosome transmission, especially if most diverted water contact is only for domestic activities. Future studies are needed to experimentally investigate the activities diverted when safe water is provided to rural communities.

Safe water provision was influenced by choice of location, even at very granular scales (within 1000m), and by control programme objectives. At fine spatial scales, predicted usage of a tap/borehole when the optimal location was chosen was 5.5 times higher than tap/borehole usage with random placement. For programmes optimising the percentage of the population with access to a tap/borehole, ideal locations were in densely populated areas without any taps/boreholes. For programmes seeking to reduce open water contact, ideal locations balanced population density and closeness to the shoreline to maximise crowding out. While a 2023 WHO NTD guideline called for multisectoral WASH-NTD collaboration [47], our results show the need to have a clear programme objective and disease-specific theories of behaviour change. Our modelbased suggestions for where infrastructure should be placed, need to be tested in future interventional research with exploration of hydrological and other location constraints for building taps/boreholes in conjunction with qualitative work to investigate user preferences and attitudes for safe water interventions. Our findings were based on existing, available infrastructure but could be used to guide the placement of new technologies if taps/boreholes are not preferred.

The spatial decay models we proposed have practical applications across different elements of multifaceted schistosomiasis control, emphasised by the WHO [27]. As essentially all open water contact and tap/borehole usage occurred within 1 km from the homes of individuals, exposure reduction interventions may need to focus on local open water contact, limited to the home village and neighbouring villages only. Our models were able to predict the number of people using each open water site, which could guide focal mollusciciding of the most used sites to reduce transmission [28]. The results suggest that mollusciciding via radius control should be executed within 1 km of households–larger than the 500m radius suggested by the WHO [28]–if the aim is to treat essentially all locally relevant sites. We found that GPS logger data from 25 individuals over ten days would have been sufficient to adequately estimate the spatial decay function, highlighting the logistical feasibility of this method. In addition, models generalised well across three diverse districts in Uganda, and can be used with publicly available building footprints or WorldPop data [48, 49] to predict population-level water usage patterns.

We extensively investigated the generalisability of our models, data, and potential limitations within our study. We focused on the spatial dynamics of water contact and did not model temporal patterns. In this study, 43% of individuals had water contact across more than two days and 16% across all days. The small sample with frequent water contact across the 10-day observation window limited efforts to analyse recurrent water usage patterns, despite our study being in the dry season when water contact is the highest [15, 24]. We do however report on water usage patterns by time of day. Our spatial decay models provide the same predictions for all household members, limiting their ability to study intra-household dynamics of water contact. While we built spatial models where the spatial decay varied based on age, gender, and occupation, we found that these models did not outperform a purely spatial model ignoring sociodemographic factors. Due to the nature of the GPS logger data, we had no information on specific activities. Activity information is better captured through surveys or direct observation [15] and has been described extensively for the SchistoTrack cohort elsewhere [24]. In the event of individuals changing their behaviour due to wearing the loggers, we removed the first day of observation from the analysis. We also did not receive any reports (though not elicited in a structured manner) that individuals changed their routine behaviours while wearing the loggers. To investigate potentially misclassified transient movement next to water sites as actual open water site or tap/borehole usage, we assessed if the spatial decay parameters b_0_ and b_1_ changed when we successively removed short duration contact events and found that our models were robust until all events <5 min were removed. Yet, this study may not have captured all local water contacts in case some open water sites were not mapped, or people travelled beyond the study area to contact sites, leading to an underestimation of water contact. The median duration of water contact of 3 min per person per day is similar to an observation study of 120,000 water contacts in an endemic area in Senegal where the duration was 4.3 min per day [50]. Another study in Kenya found that people typically visited <1 open water site, in line with our findings showing visits mostly to one site [26]. Our study mapped open water sites up to 26 km away, and as >99.5% of all water contact occurred with 3000m, it is unlikely that mapping further-way sites would have changed the findings. We also note that our study focused on communities within 2 km of waterbodies. Our findings may thus not apply to communities further away from the shoreline.

In conclusion, we provide generalisable individual-level spatial decay models of human water usage with a focus on schistosomiasis. Future research should investigate to which extent similar models could be applied to other waterborne diseases such as diarrhoea or cholera. There is a need for GPS logger data collection across different geographical settings to establish the generalisability of the observed spatial decays. With cross-setting validations, our models could be used as a planning tool for control interventions such as WASH and mollusciciding or to prioritise areas for MDA.

## 4 Methods

### 4.1 Study setting and participant sampling

The GPS logger study was conducted as part of the SchistoTrack cohort study baseline in January and February 2022 across Pakwach, Buliisa, and Mayuge districts in Uganda [29]. Twelve villages were selected among the original 38 villages in Schisto-Track cohort. In each district, four villages were purposively sampled based on their levels of water contact and occupational fishing, determined predominantly by the presence of a beach or landing site. None of the selected villages had piped water. A maximum of 50 participants per village were recruited, with a target of one adult-child pair (adult [>18 years] and one child [5–17 years]) per household. Households were selected among SchistoTrack study participants (n = 2885) who had been recruited via random sampling from village registers or MDA records. For this nested crosssectional study, SchistoTrack participants were sampled to ensure a balance of age and gender, and fishermen were oversampled to ensure a diverse representation of water contact and mobility patterns. We excluded participants who were physically immobile as defined by the participant, and thus unable to have water contact. We also excluded children who were sent away to boarding school and unavailable during the GPS logger study period. All participants were unaware of their schistosome infection status when they entered the GPS logger study.

### 4.2 Community sensitisation, consent, and remuneration

Goylette F. Chami (G.F.C.), Max T. Eyre (M.T.E.), and Fabian Reitzug (F.R.) conducted workshops in each village to explain the study objectives and address any questions or concerns. As part of the standard written informed consent for recruitment into SchistoTrack, consent was obtained from adult participants and parents or guardians for children under 18 years in the local language specifically for the GPS logger study. Children provided verbal assent and, where possible, written assent was also collected. Participants received a nominal remuneration equivalent to about one day of wages (UGX 10,000, approximately USD∼ $2.73) in addition to any incentive in the study upon returning their GPS loggers at the end of the observation period.

### 4.3 GPS logger settings and procedures

Wearable GPS loggers (i-gotU GT-600; Mobile Action Technology, Taiwan) were used to record the location data of participants at two-minute intervals between 5 a.m. and 8 p.m. local time over approximately ten consecutive days. This time period was chosen to capture activities during the day. Loggers were turned off from 8 p.m. to 5 a.m. to save battery using scheduled control, as loggers were not recharged in the field by participants. All loggers were set to motion activation such that loggers turned off when participants did not move—also to save battery. Button control on devices was disabled such that participants were unable to switch them off. Devices were placed in waterproof pouches and worn around the neck using a lanyard (Fig. S9). Children and adults within the same household always received different coloured lanyards to minimise the risk of accidental mixing up of the loggers. Adults were asked to help children with the day-to-day wear of the loggers. Participants were instructed to wear the devices during all daily activities, including during water contact, and to remove them only at night.

### 4.4 Participant flow

A participant flowchart is shown in Fig. S10. Two hundred and fifteen loggers were purchased for the study with the aim to hand out 200 loggers per district, i.e., 600 in total. A total of 585 participants were recruited, slightly below the target of 600. This was due mainly to device failure and loggers not being returned by participants after hand out in Pakwach. After excluding participants with fewer than two complete days of GPS data—excluding the first day of recording for all participants to reduce Hawthorne effects [51]—we included a final sample of 452 participants in the study.

### 4.5 GPS logger testing and validation

We tested the accuracy of GPS loggers in situ. During fieldwork, ten loggers were put next to each other on the ground for 10 minutes in an unobstructed area in Buliisa to estimate GPS position error in the field. Based on this experiment, we estimated that GPS loggers were accurate within 8.6m. GPS precision was estimated using the uere function in the ctmm package in R [52]. Our estimate of 8.6m aligned with a previous study which estimated the accuracy of i-gotU GT-120 under different levels of obstruction and found that the mean location error was less than 10m and that significant cover moderately increased location error by 2.2m [53].

### 4.6 GPS logger data retrieval

For 94 participants, no GPS data could be retrieved as loggers suffered from water damage despite the use of protective pouches and indication from the manufacturer, Mobile Action, that the devices were fully waterproof. Due to corrosion of the metal contacts, these loggers could not be pulled using a computer, as they were not recognised by the i-gotU GPS logger software. We undertook multiple steps to retrieve data from the affected loggers. We cleaned the metal contacts with ethanol and attempted to retrieve the data in the field. We transported all loggers to Oxford, cleaned them again with ethanol, and attempted to retrieve the data once more. At that point, there were still 36 GPS loggers with missing data; we opened their cases at the University of Oxford Materials Science Department, where a trained research technician resoldered the GPS logger chips onto a functioning GPS logger. This enabled us to retrieve data from 24/36 GPS loggers.

### 4.7 Self-reported wear compliance

When participants returned GPS loggers, we asked about wear comfort of the loggers. As not all devices were returned immediately following the end of the study period by the participants themselves, this data was available for 87% of participants (395/452). Among those participants, 83% (328/396) reported that wearing the logger was comfortable. Participants self-reported loggers causing perceived chest pain (34), body weakness (5), nausea (1), or headaches (1), as reasons for discomfort, which were addressed through sensitisation.

### 4.8 Deriving open water contact and tap/borehole usage patterns

As part of the study, F.R., M.T.E., and initially G.F.C., comprehensively mapped the locations of all taps/boreholes used by the study communities, assisted by the village chairman or a village health team member. Trained malacologists from the Division of Vector-Borne and Neglected Tropical Diseases, Uganda Ministry of Health, mapped all open water sites. A contact event with an open water site and tap/borehole was defined as being within a 30m buffer of the GPS location of an open water site or tap/borehole. We chose 30m to account for possible GPS position errors while minimising overestimation of water contact. Past GPS logger studies have used similar-sized or larger buffers. Eyre et al. used a 30m buffer around the shoreline while Seto et al. used a 100m buffer around the shoreline [33, 39]. GPS locations of sites and taps/boreholes were recorded via tablets (Lenovo Tab M8 (3rd Gen); Lenovo Group Limited, China) with 5–10m manufacturer-reported accuracy and GPS logger points with 8.6m accuracy, as reported above. All contact events with open water sites and taps/boreholes were identified using recursion analysis, which is a well-established method in movement ecology [54, 55] and which was implemented in the recurse package in R [56]. The output from the recursion analysis was a list of all distinct open water contact and tap/borehole usage events with an associated timestamp, number of revisits, contact duration, and open water site and tap/borehole identifier. Compared to simply counting the number of points inside each buffer area, recursion analysis has the advantage that it reconstructs an approximated, linear trajectory and is therefore able to identify visitations with durations less than the GPS logger sampling frequency. This is particularly important given that water contacts are typically short [50].

As the SchistoTrack protocol did not allow sites to be larger than 15m, even when they were directly adjacent, we grouped such nearby locations into a single open water site or tap/borehole for the analysis, following methods used in Iacovidou et al. [30]. The density-based spatial clustering of applications with noise (DBSCAN) algorithm was used and the optimal number of clusters was determined based on the gap statistic, separately by district [57]. This collapsed the 143 water sites and the 63 taps/boreholes into 69 and 32 clusters, respectively, to which we then assigned the mean GPS cluster coordinates.

### 4.9 Sanitation infrastructure

This analysis did not focus on the use of public latrines, even though it would have been possible to derive this information from the GPS data, using the same methods as for open water site and tap/borehole usage. There were two reasons for this. First, field visits to all public latrines by M.T.E. and F.R., and initially G.F.C., revealed that most latrines were locked (usually a community member held the keys), and they were not regularly used by the community. Second, among the 452 GPS logger participants, 343 (75.9%) lived in households that had a private latrine (i.e., a flush or pour-flush toilet, a covered pit latrine with/without privacy, or a composting toilet). Thus, most households could rely on private latrines which provide more privacy and convenience compared to public latrines [58, 59]. However, private latrine usage events could not be derived from GPS logger data due to unknown precise location of these latrines.

### 4.10 Dataset and variables

#### Dataset

The dataset used to estimate spatial decay models was generated by calculating the Euclidean household distance from each household to all open water sites and taps/boreholes (both used and unused) within the same district (Fig. 3). This covered distances of up to 11 km, 17 km and 26 km for Pakwach, Buliisa, and Mayuge, respectively.

#### Outcomes

For the models estimating any usage, the outcome was a binary indicator of whether an individual used an open water site or tap/borehole. For the model estimating duration of usage, the outcome was a continuous variable indicating the number of minutes per day each individual had contact with each mapped open water site or tap/borehole. The duration variable was generated by dividing the total duration of contact with each open water site or tap/borehole by the number of distinct calendar days with GPS data for each individual.

#### Covariates

The main predictor variable was household distance to each open water site or tap/borehole. We selected additional predictor variables based on their relevance to water contact patterns. The following individual-level variables were used: age, gender, occupation, self-reported water contact, activity match and mobility.

Age was coded as a binary variable (< 15 years, ≥ 15 years). This cut-off was chosen because we previously showed that water contact levels were low in individuals aged < 15 years and increased substantially thereafter, which suggested 15 years of age as a natural cutoff [24]. We also used gender (male/female) reported by the household head as a covariate. Individual-level occupation was also used as a covariate because occupation is an important determinant of water contact [24]. Occupation of the household head (aged ≥ 18 years) was recorded. The occupation categories were fishing, fishmongering, farming, and other. For all participants aged < 18 occupation was coded as “none”. A self-reported water contact variable, described in detail in Reitzug et al. [24] was also included. A binary activity match variable was generated for each individual–water site pairing. Whenever an individual conducted any of the 11 domestic, recreational or occupational open water contact activities, and whenever the same activity was reported to be done at a specific site, based on reports by the local guide, this was defined as an activity match. The 11 water contact activities included getting drinking water, washing clothes with soap, washing clothes without soap, washing jerry cans or household items, washing clothes with soap, collecting papyrus, fishing, fishmongering, collecting shells, and swimming or playing. All individuals who reported getting drinking water were also assigned to having an activity match with taps/boreholes as this activity was amenable to being conducted at a tap/borehole. Apart from the activity match, water contact activities of an individual were not considered in this analysis as the GPS data does not contain any definitive information on the specific water contact activity an individual engaged in. We also used an individual’s mobility tercile as a covariate (see section below). At the household level, aside from distance to the closest sites and taps/boreholes, a self-reported variable for whether the household used a safe drinking water source was also used. Taps/boreholes were counted as safe whereas open freshwater sources such as swamps or lakes were counted as unsafe. To account for geographic, cultural, behavioural, and environmental factors that were different between study locations, a district-level categorical variable was included in some models. Households paid different prices for tap usage compared to boreholes (see results), therefore we also generated a variable indicating whether each public water supply was a tap or a borehole.

#### Schistosome (re-)infection

To test associations between water contact and (re-)infection intensity, we relied on schistosome infection data from the Schisto-Track study. All participants were tested for schistosome infection using Kato-Katz microscopy at baseline and one-year follow-up and all were treated with praziquantel, irrespective of infection status, and remained unaware of their infection status. Baseline testing took place prior to GPS logger handout and study team was unaware of Kato-Katz results. Infection intensity was measured in eggs per gram of stool. One-year post-treatment infection measurement was chosen as this is the interval for MDA in endemic areas and frequently used as a time-frame to assess reinfection [27, 60, 61]. Among participants, 20% (91/452) were missing Kato-Katz results at one-year follow up. For those individuals, we imputed reinfection intensity based on baseline infection intensity using multivariate imputations by chained equations, implemented in the mice package in R [62].

#### Geospatial data

All waterbody boundaries are from a public shapefile of waterbodies in Uganda, made available online under a Creative Commons Attribution (Non-commercial 4.0 International) by the World Resources Institute [63]. Building footprints were obtained from the Global Google–Microsoft Open Buildings Dataset (V3), which has mapped over two billion structures globally and is made available under a Creative Commons Attribution (4.0) licence [48].

### 4.11 Human mobility measures

We aimed to compare mobility patterns across different activity types: visits to open water sites, taps/boreholes, and overall movement. To do so, we selected the radius of gyration R*_g,i_* as our metric, as it measures the Euclidean typical displacement of individual i from their household location. This choice ensured consistency with our use of Euclidean distances for calculating household distances to visited open water sites and taps/boreholes. R*_g,i_* was computed using the following formula:

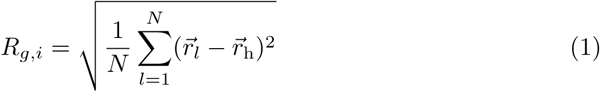

where 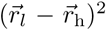 represents the squared Euclidean distance from location l to the household location h.

### 4.12 Spatial decay models

The aim of this analysis was to model open water site usage and tap/borehole contact behaviour of an individual based on Euclidean household distance to open water sites and taps/boreholes. This approach was motivated by evidence that water contact and schistosome infection are waterbody distance-dependent [3, 24, 26]. Our previous work using self-reported water contact and closest site assignment suggested that the water contact over household distance was approximately linear [24]. The modelling framework proposed here differs from previous studies in that it aims to estimate usage of each open water site and tap/borehole, as opposed to just the closest site. Our spatial decay models take the general form:

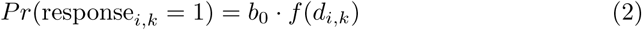

where Pr is the probability that individual i uses a specific water site and tap/borehole k. Here, f(d*_i,k_*) represents the spatial decay function with d being the household distance to k. The binary outcome P is modelled using a Bernoulli distribution with an identity link function to represent probabilities directly. All spatial decay models were fitted as Bayesian non-linear regression models on a training dataset containing 85% of the full data and evaluated by calculating the area under the receiver operating curve (auROC) on 15% held-out data.

The model fitting process was as follows. First, we determined a suitable spatial decay function (f(d)) in eq. 2. We tested whether an exponential decay, a power law decay—two commonly used decay functions in human mobility modelling [32]— or a Hill function best fit the data. As models were fitted as Bayesian non-linear models, the best decay function was determined via ELPD —a Bayesian leave-one-out measure of out-of-sample predictive fit [64]. Second, we tested whether the inclusion of covariates, adding one j variable in eq. 3 at a time (district, gender, age (<15 years, ≥ 15 years), occupation category, or improved household drinking water source) improved model fit compared to the base model without additional covariates (baseline model, see eq. 2).

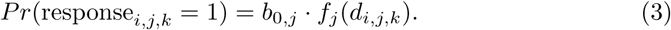

We modified spatial decay models described in eqs. 2–3 to evaluate the influence of spatial distance, human mobility, and usage of safe water infrastructure on human contact with open water sites, as shown in Fig. 3. The model to assess the influence of household distance on open water contact (Fig. 3a), is shown in eq. 3. Our approach to model the influence of having access to different, competing water sites, on water site usage (competing opportunity hypothesis in Fig. 3b) borrows from Stouffer’s intervening opportunity model, where aggregate mobility between cities is driven by the number of opportunities at the destination versus in-between origin and destination [65]. We develop a spatially explicit competing opportunity model where the spatial decay parameter b_1_ is estimated separately for the first, second, third, . . . , n*^th^* closest site k_1_*_−n_* in eq. 3. As <1% (4/452) of individuals visited more than a five different open water sites, we collapsed all sites beyond the 5^th^ closest into one category. We assessed the role of human mobility (Fig. 3c) by estimating b_1_ separately by for each tercile of mobility, represented by j in eq. 3. We assessed the role of taps/boreholes on open water contact by constructing a crowding out model (see Fig. 3d and eq. 5). Analogously to the model predicting the probability of open water site usage in eq. 2, we used the same modelling approach to predict tap/borehole usage, with P(X = 1) representing the probability of using a tap/borehole. We also used spatial decay models to predict the duration of open water site and tap/borehole usage. These models took the following form:

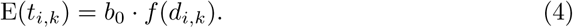

This model is a negative binomial model to account for overdispersion, where

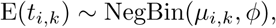

with µ*_i,k_*= b_0_ · f(d*_i,k_*) as the mean duration, and ϕ as the dispersion parameter, capturing variability beyond what is expected under a Poisson model.

Extreme duration values can lead to exaggerated values of the dispersion parameter ϕ, making the model appear more overdispersed. Thus, all duration values > 99.9^th^ percentile, corresponding to a duration of > 123 minutes per day, were capped by setting these values to 123 minutes. For exponential duration decay models, we used an uninformative prior Unif(0, 1) for b_1_ and a normal prior Normal(1, 9), truncated at zero and 30, for b_0_. This prior was chosen to match the mean and standard deviation of the data.

We also built spatial decay models in which usage of taps/boreholes reduces the probability and frequency of open water site usage (crowding out models), and compared predictive performance to models without crowding-out effects. For the probability of open water site usage with crowding out, the model took the following form:

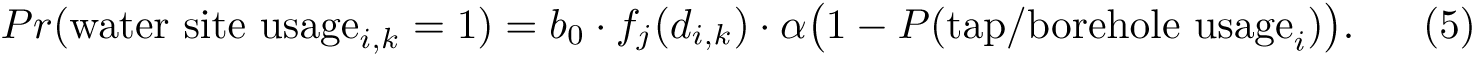

Here, the probability of open water site usage is inversely related to the probability of tap/borehole usage and α quantifies the magnitude of crowding out. (An analogous model is used for the duration of water site usage.)

### 4.13 Estimating spatial decay models

We used Bayesian non-linear regression models implemented using the brms package [66] in R version 4.1.0 to fit spatial decay models. Bayesian modelling was used because it allowed us to estimate distributions of the decay parameters and quantify the uncertainty of these parameter estimates by taking posterior draws from these distributions to obtain 95% credible intervals. Due to convergence issues in the Bayesian modelling, we restricted our data to all sites within 3000m of the households, which captured over 99.6% of all water sites and taps/boreholes used. We used district-specific intercepts across all models to account for geographic differences in baseline levels of water site and tap/borehole usage. As only seven individuals lived within < 20m distance of water sites or taps/boreholes, predicted values were reported for distances between 20–3000m. For all Bayesian models estimated via brms, a Hamiltonian Monte Carlo sampling algorithm via the Stan backend, employing the No-U-Turn Sampler (NUTS), was used [66, 67]. Four Markov chains with 2000 iterations per chain (1000 warm-up and 1000 sampling iterations) were employed. Convergence was assessed based on whether the potential scale reduction factor (R^^^) was close to 1.

Within brms, the plot function was used to visually assess chain mixing, and the pp check function was used to assess agreement between predicted and observed response values.

As far as possible, we used non-informative priors. For instance, in the exponential decay model, we used uniform priors Unif(0, 1) for b_0_ and b_1_, the most conservative priors possible as b_0_ and b_1_ can only take values between 0–1. For all models predicting the probability of usage, we ensure that P(response*_i,j,k_* = 1) remains a valid probability (i.e., within the range [0, 1]). To this end, the product b_0_ · f(d*_i,k_*) is constrained such that 0 ≤ b_0*,j*_ · f*_j_*(d*_i,j,k_*) ≤ 1. This condition is enforced during model fitting.

All Bayesian models (except for those where entire districts were held out, see below) were fitted on a training dataset containing 85% of the full data. For all models predicting the probability of water usage, we converted outputted probabilities between 0 and 1 into a binary outcome that could be compared with the true outcome data The optimal cut-point probability was determined using the cutpointr package, with the F1 score as the criterion [68].

### 4.14 Selecting the best fitting models

To determine which decay function best fit the data, we used Bayesian leave-one-out cross-validation, and the best fit was identified based on the expected log pointwise predictive density (ELPD) for a new dataset—a Bayesian leave-one-out measure of out-of-sample predictive fit [64]. For the decay function, ELPD was compared between global Hill, exponential, and power-law decay models. Whenever we compared whether any of the decay models depicted in Fig. 3b–c yielded improved performance over the baseline spatial decay model (Fig. 3a) or logistic regression models, we used auROCs calculated on 15% held-out data to evaluate performance. In models where one district at a time was held out to evaluate performance on the other districts (Fig. S7), the training data consisted of 150 individuals sampled with replacement from the full data to ensure the size of the training data was balanced across districts.

Importantly, we evaluated predictive performance of the spatial decay models at the individual level which is substantially more stringent compared to how gravity models are conventionally evaluated. As gravity models are not individual-level, Pearson correlation coefficients or R^2^ between predicted and observed mobility flows are typically used to evaluate gravity models [69–71]. For fine-scale predictions gravity models have performed poorly with a R^2^ values for predicting commuting behaviour in London between 7–22% [70]. We also report R^2^ here (and achieve values of 54% for open water sites), but we are not primarily interested in measures of aggregate performance but in the more difficult problem of whether individual-level usage patterns can be correctly predicted. This problem is more difficult because, for instance, if an individual visits a different water site than the one predicted, this introduces two classification mistakes from the perspective of individual-level performance evaluation while the more lenient criterion of whether this individual has water contact is still satisfied.

### 4.15 Out-of-sample prediction

The ability of spatial decay models to predict population-wide patterns of use of field-recorded water sites was demonstrated using the publicly available Global Google– Microsoft Open Buildings Dataset (V3), which has mapped over two billion structures globally [48]. Footprints of all buildings in a 10 km × 10 km area around the study villages in Pakwach were extracted. As there were few commercial or public buildings in this rural area, we assume that each building represents a household.

To generate population-wide predictions of water contact, we calculated the centroid location of each building and input the centroid GPS coordinates into eq. 3, which had been trained on the GPS logger data. This produced out-of-sample predictions of water site usage for all households in the study villages in Pakwach.

### 4.16 Simulations to identify optimal borehole locations

To suggest optimal locations for building taps/boreholes within the study area, defined here as a convex hull around the household locations with a buffer of 2000m, we used a simulation approach with previously trained models predicting tap/borehole usage for strategy i and crowding out for strategy ii. We divided the study area into a 400m × 400m grid and iterated over each cell, placing one borehole at the centre of the index cell *i* and obtaining fitted values for tap/borehole usage probability across all cells (outcome for strategy i) or duration of open water site usage (outcome for strategy ii). We then summed the values across all cells, weighted by population density based on building footprints (see above), and assigned them to cell *i*. We defined the optimal locations as the cell with the largest value for strategy i as this maximised tap/borehole usage and the smallest value for strategy ii as this maximised crowding out of open water contact, respectively. We further ranked all cells from 0 (worst) to 1 (best) to enable a direct comparison of how each cell was ranked based on strategy i and ii).

### 4.17 Sensitivity and robustness analyses

Several sensitivity and robustness analyses were conducted to account for the potential of water contact misclassification, systematic differences in compliance across demographic groups, and differential logger failure across districts.

As most water contact events are brief in duration (in terms of minutes), there is a risk of misclassification, as such water contact may represent a different type of activity, for instance passing by a water site without having actual water contact. To evaluate the influence of such brief, potentially misclassified contact events, we successively increased the threshold below which we removed events (0–10 min). We then recalculated the spatial decay model predicting water site usage including only water contact events above these thresholds to understand how model coefficients b_0_ and b_1_ changed. This thresholding exercise also informed on the minimum GPS logger sampling frequency needed to reproduce the patterns observed with GPS data at two-minute intervals.

Comparisons were made between the demographic characteristics of individuals with sufficient GPS data (≥ 2 days), those with insufficient data (< 2 days, or completely missing data), and the broader SchistoTrack study population. This analysis provided insights into potential selection biases arising from sampling/distribution of loggers and participant non-compliance. Among those with GPS data, the number of GPS points per individual per day was used as an indicator to examine differences in wear non-compliance between demographic groups.

This study was conducted across three different districts, and behaviour patterns from one district may not be generalisable. To address this, data from two districts at a time were used to predict water site usage in the held-out district.

The number of individuals and observation days were down-sampled to determine the minimum number of individuals and observation days required to achieve good model performance.

## Ethical approval declarations

Data collection and use were reviewed and approved by the Oxford Tropical Research Ethics Committee (OxTREC 509-21), Vector Control Division Research Ethics Committee of the Uganda Ministry of Health (VCDREC146), and Uganda National Council of Science and Technology (UNCST HS 1664ES). Written informed consent was obtained for all study participants with adults consenting on behalf of children and children providing verbal assent.

## Supplementary information

This file contains supplementary text, figures, and tables.

## Acknowledgements

We are thankful for the involvement of our study participants and local communities. We thank all field teams, including malacologists, surveyors, nurses, and laboratory technicians involved in the 2022 SchistoTrack baseline data collection, especially Benjamin Tinkitina, who led the household survey team. The support of the Uganda Ministry of Health, local district leaders, focal health workers, and village health teams was crucial for building partnerships and continued trust with study communities. We thank Farina L. Shabaan for help with preparing the GPS loggers for fieldwork. Post-fieldwork, Adam Fowler helped with initial attempts to retrieve data from damaged GPS loggers. Lauren B. Wilburn, Keiran Foster, and Joseph Brown pulled GPS logger data from damaged GPS loggers. Prathyush Sambaturu gave feedback during the initial stages of the analysis.

## Declarations

### Code availability

The computer code used to provide all figures and tables in the main manuscript and supplementary materials will be made available on FigShare upon publication.

### Data availability

Due to the highly sensitive nature of the GPS logger data, restrictions from the data protection impact assessment, and ongoing nature of the SchistoTrack cohort, these data are not publicly available. Extensive metadata for all variables have been provided in the manuscript and supplement.

### Author contributions

Conceptualisation: F.R. and G.F.C. Data curation: F.R., N.B.K., B.N., M.T.E., and G.F.C. Formal analysis: F.R. Funding acquisition: G.F.C. Investigation: F.R. and G.F.C. Methodology: F.R. and G.F.C. Project administration: N.B.K. and G.F.C. Resources: N.B.K. and G.F.C. Software: G.F.C. Supervision: G.F.C. Validation: F.R. Visualisation: F.R. Writing – original draft: F.R. Writing – review & editing: F.R., N.B.K., B.N., M.T.E., and G.F.C.

### Open access

For the purpose of Open Access, the author has applied a CC-BY public copyright licence to any Author Accepted Manuscript version arising from this submission.

## Supplementary information

## Appendix A Supplementary text

### A. 1Additional validation and robustness analyses

The minimal number of individuals and observation days, i.e. the minimum data, needed for the spatial decay models was established by holding out entire individuals, GPS logger days, and districts. With all individuals included, performance plateaued after just 15 individuals were used to train the decay model (Fig. S7a). Using a random subset of all days (1–6 days), one day of data was sufficient to achieve model performance similar to using all six days when predicting water site usage (Fig. S7b). Holding out entire districts revealed that the data from Buliisa were best able to predict the decay function for the two held-out districts (auROC = 0.910, 95% CI: 0.905–0.916). As the decay functions of Mayuge and Pakwach were more similar (Fig. S11), training the model on Mayuge performed best at predicting Pakwach, and vice versa (Fig. S7c). We also assessed the impact of GPS logger sampling frequency (2 min) on the parameter estimates in the global spatial decay model predicting water site usage. To this end, we successively removed all open water site usage events of less than the cut-off *c* for *c* ∈ {0, . . . , 10}. By estimating b_0_ and b_1_ for the restricted dataset based on *c*, we obtained the minimum sampling frequency needed to obtain parameters within the 95% CrIs we observed with the full data. The results are shown in Fig. S8.

Sufficient data from GPS loggers were retrieved for 452/584 participants who were offered wearable GPS loggers. To investigate differences between individuals with and without missing data, their demographic characteristics were compared in Table S4. Additionally, comparisons were made between individuals selected for the GPS logger study (n = 584) and SchistoTrack participants from the same villages who were not selected for the GPS logger study (n = 379). There were differences regarding district and gender, as gender composition and missingness varied by district. No significant differences were observed between included and excluded participants regarding age, self-reported water contact, occupation, or distance to the nearest water site (Table S4). Most importantly, our model predictions did not display obvious biases in prediction errors. Socio-demographic characteristics of individuals whose water site usage was correctly predicted were similar to those whose patterns were misclassified (Table S6).

As the mean number of GPS observation days varied by district (Pakwach = 13, Buliisa = 12, Mayuge = 11), wear non-compliance was investigated using district-specific terciles of the number of GPS points per person per day (Table S5). This allowed us to understand whether relative levels of compliance varied based on sociodemographic characteristics. We found no significant differences across any of the included variables, except for age. The proportion of those aged ≥ 15 years was higher (64.7%) in the tercile with the highest number of points compared to the bottom tercile (51.0%, p = 0.04), indicating that adults may have been more compliant. There was no evidence that device failure due to water contact introduced information bias as the number of observation days was similar between participants with and without GPS-derived water contact (11.2 versus 11.0 days, p = 0.72).

We found that the spatial decay model with district-specific intercept outperformed all other decay models in Table S3, including those with different decays based on socio-demographic, behavioural and WASH factors. Nonetheless we wanted to understand if there were any systematic biases in the ability of the spatial decay model in predicting open water site usage across demographic groups. To this end, we compared differences between individuals <25^th^ percentile of predictive accuracy, 25^th^–75^th^, and ≥ 75^th^ percentile (Table S6. Based on this comparison, there were no significant differences across age and gender (as both p > 0.05). There were significant differences regarding occupation as fishermen were somewhat more represented in the lower 25^th^ percentile of accuracy compared to the other groups (16.4% versus 8.1% and 5.3%,p < 0.01). At the same time, a spatial decay including occupation did not outperform the district-specific decay model (Fig. S3). The results in Table S6 also reveal that predictions were less accurate for people in households using improved drinking water sources (p < 0.01). There were no differences in predictive accuracy based on mobility. Distance also influenced predictive accuracy as household of people in the lowest quartile were a median of 95.5m and 187.8m away from open water sites and taps/boreholes respectively while these same statistics were 376.4m and 444.4m for people in the top quartile (both p < 0.01).

## Appendix B Supplementary figures

**Fig. S1:**
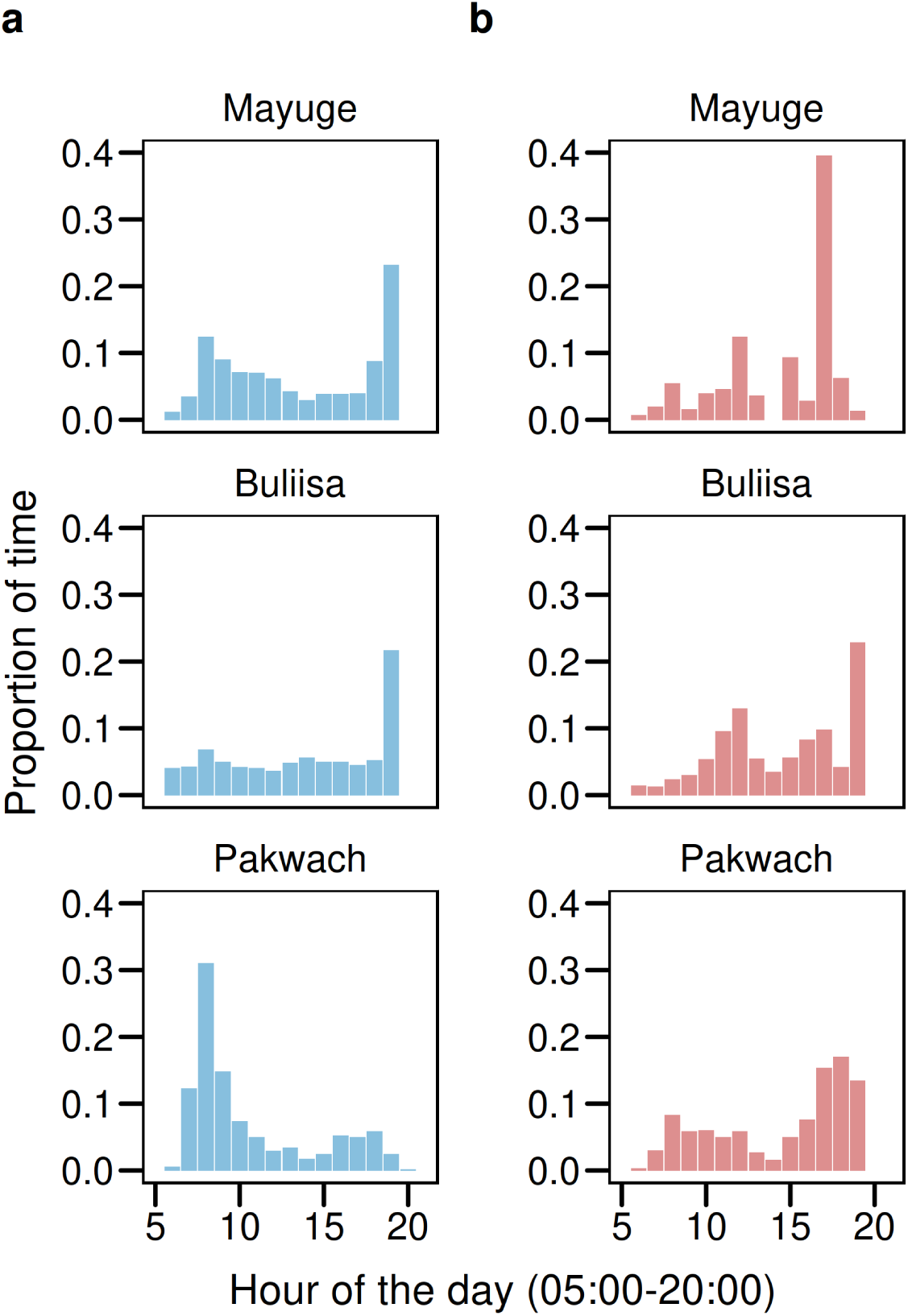
Diurnal patterns of open water site and tap/borehole usage. **a** Open water site usage by hour of the day, shown for GPS logger observation hours (5 a.m.–8 p.m.). **b** Tap/borehole usage by hour of the day.

**Fig. S2:**
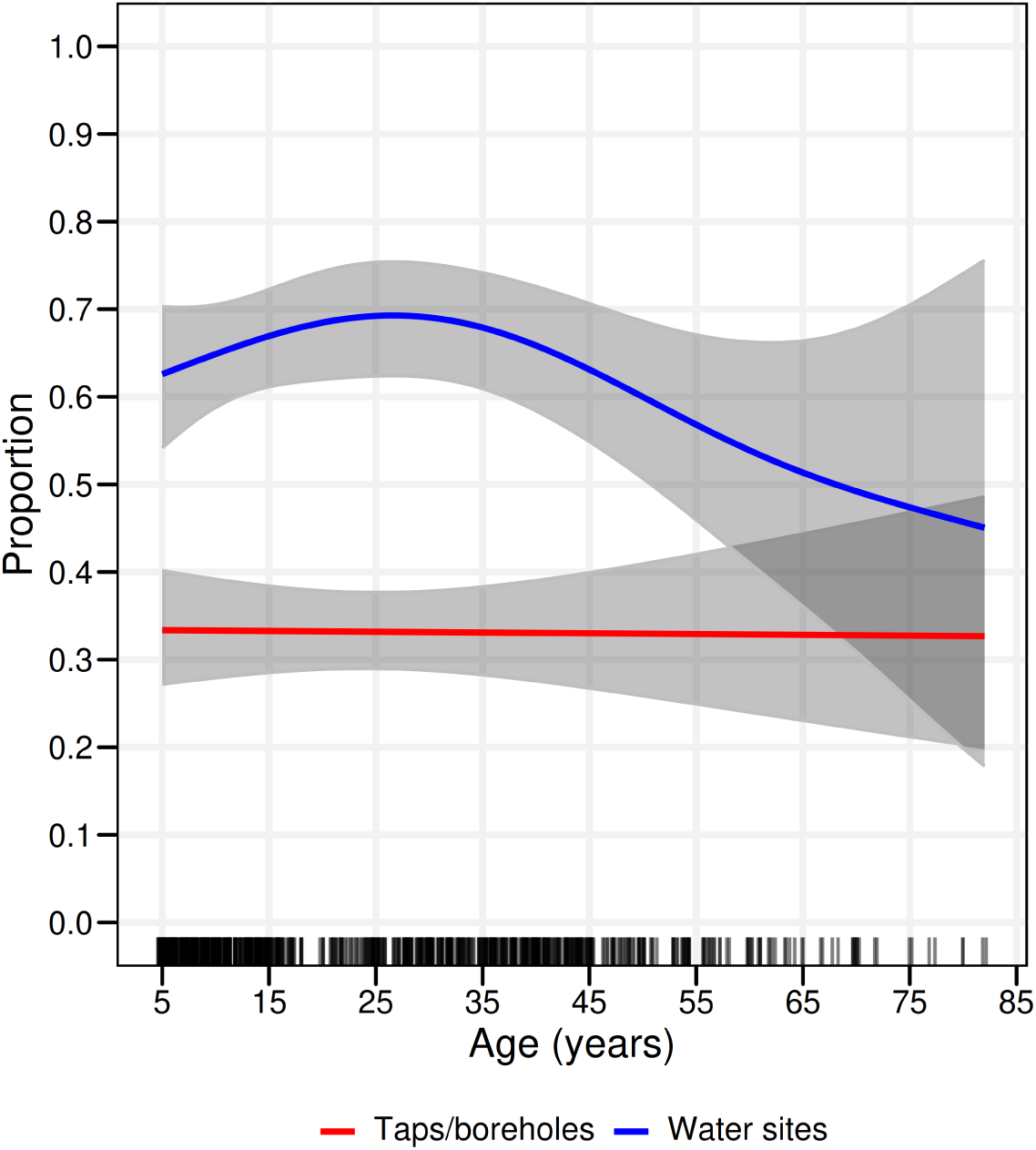
Open water site and tap/borehole usage over age. Generalised additive models showing the proportion of individuals using open water sites and taps/boreholes over age. Shaded grey areas represent 95% confidence intervals.

**Fig. S3:**
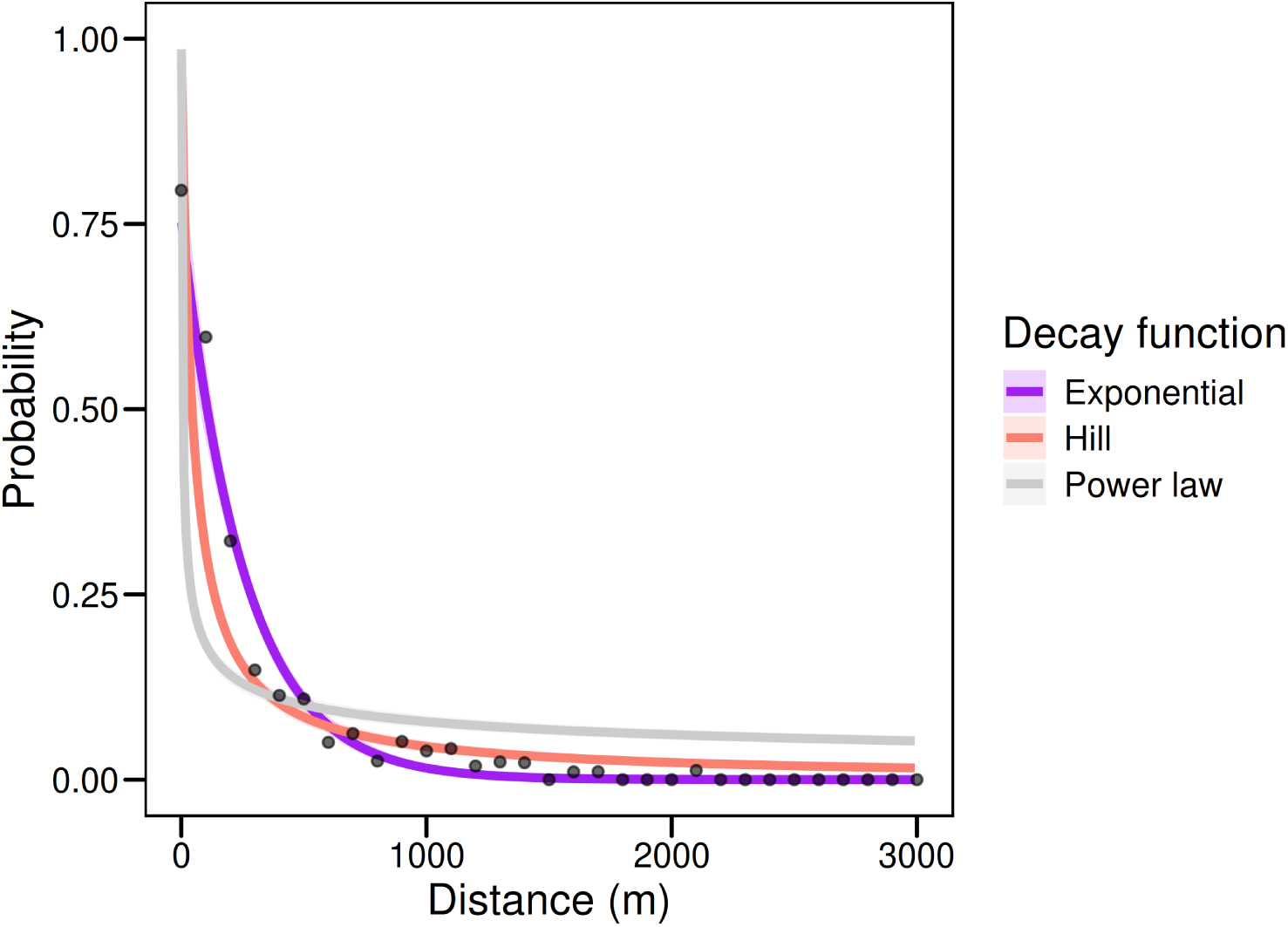
Comparison between different spatial decay functions. Estimated spatial decays from Bayesian models using a power law decay, exponential decay, and a Hill function. A model comparison based on expected log pointwise predictive density difference suggested that the exponential decay outperformed both the power and Hill functions (p < 0.01). Black dots represent the true proportion of individuals using open water sites over household distance with values plotted in 100m increments. Shaded grey areas represent 95% credible intervals.

**Fig. S4:**
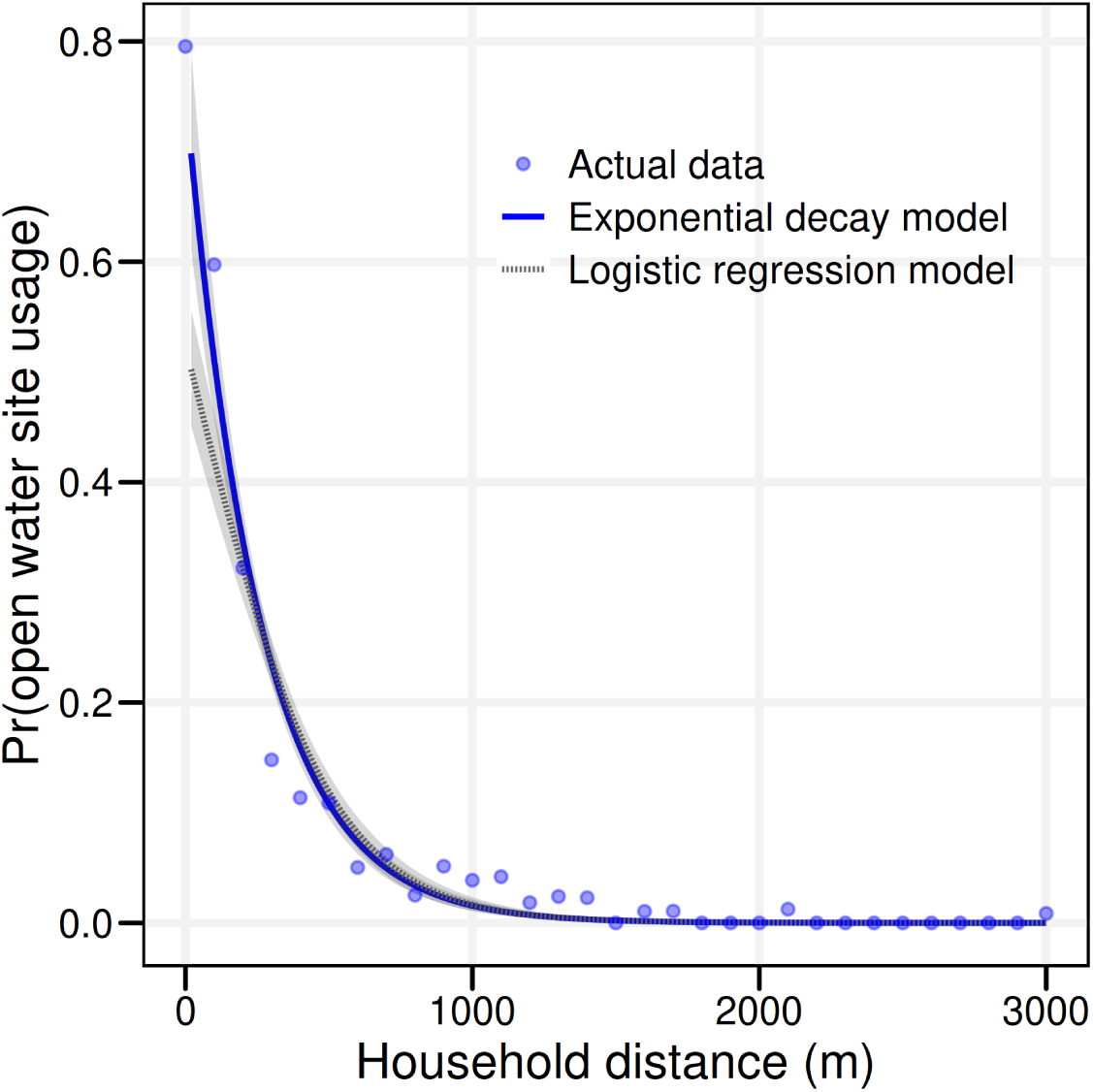
Comparison between exponential spatial decay and logistic regression models. Lines and shaded areas represent predicted values and 95% credible intervals of predicted open water site usage probability from exponential spatial decay and logistic regression models, respectively. Blue dots represent the true proportion of individuals using open water sites over household distance with values plotted in 100m increments. The exponential spatial decay model outperformed the logistic regression models based on expected log pointwise predictive density difference (p < 0.01). Shaded grey areas represent 95% credible intervals.

**Fig. S5:**
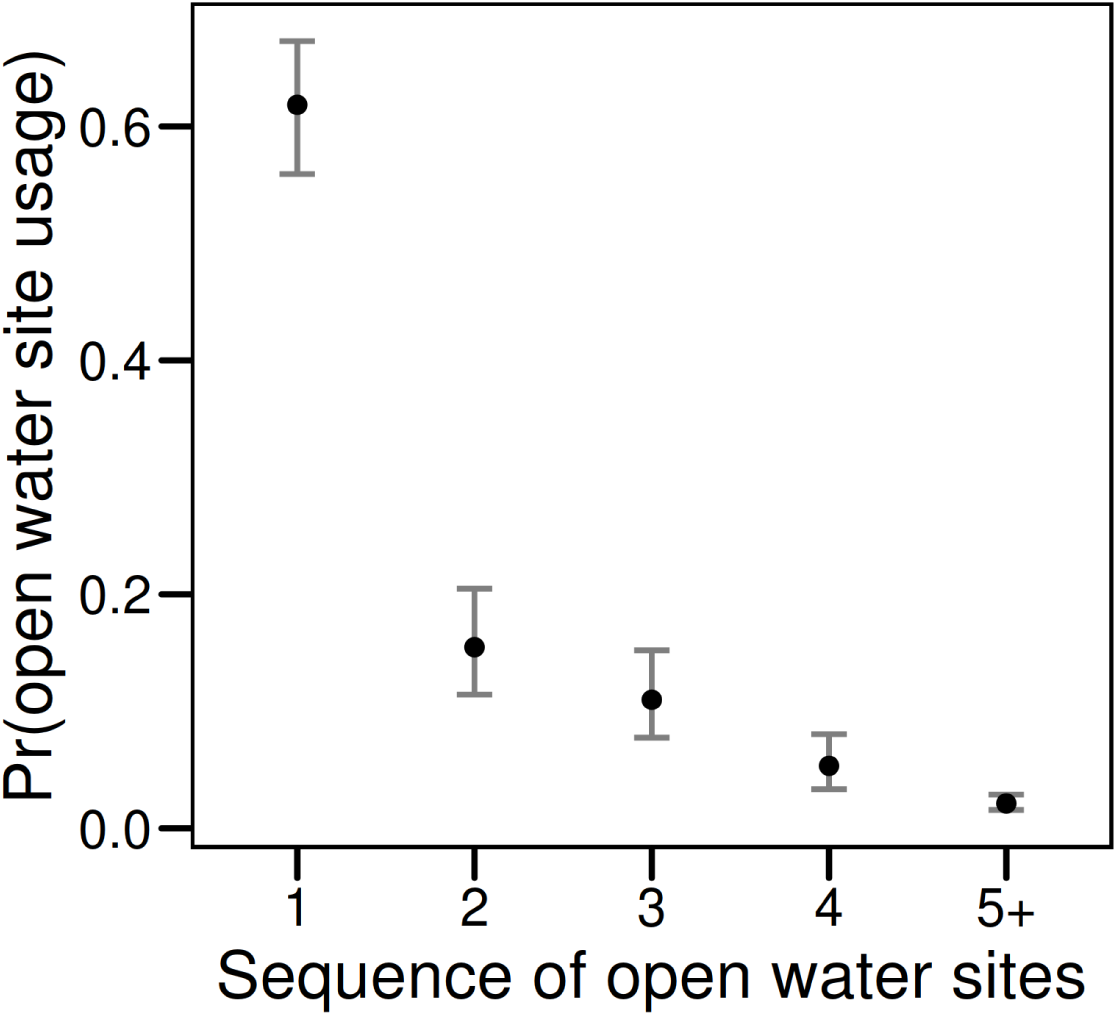
Open water site usage by sequence of sites. Logistic regression model predicting open water site usage probability of an individual by the sequence of sites, that is based on whether a given site is the closest, second closest … fifth closest/further away site. Black dots represent point estimates, error bars represent 95% credible intervals. Vertical bars represent 95% credible intervals.

**Fig. S6:**
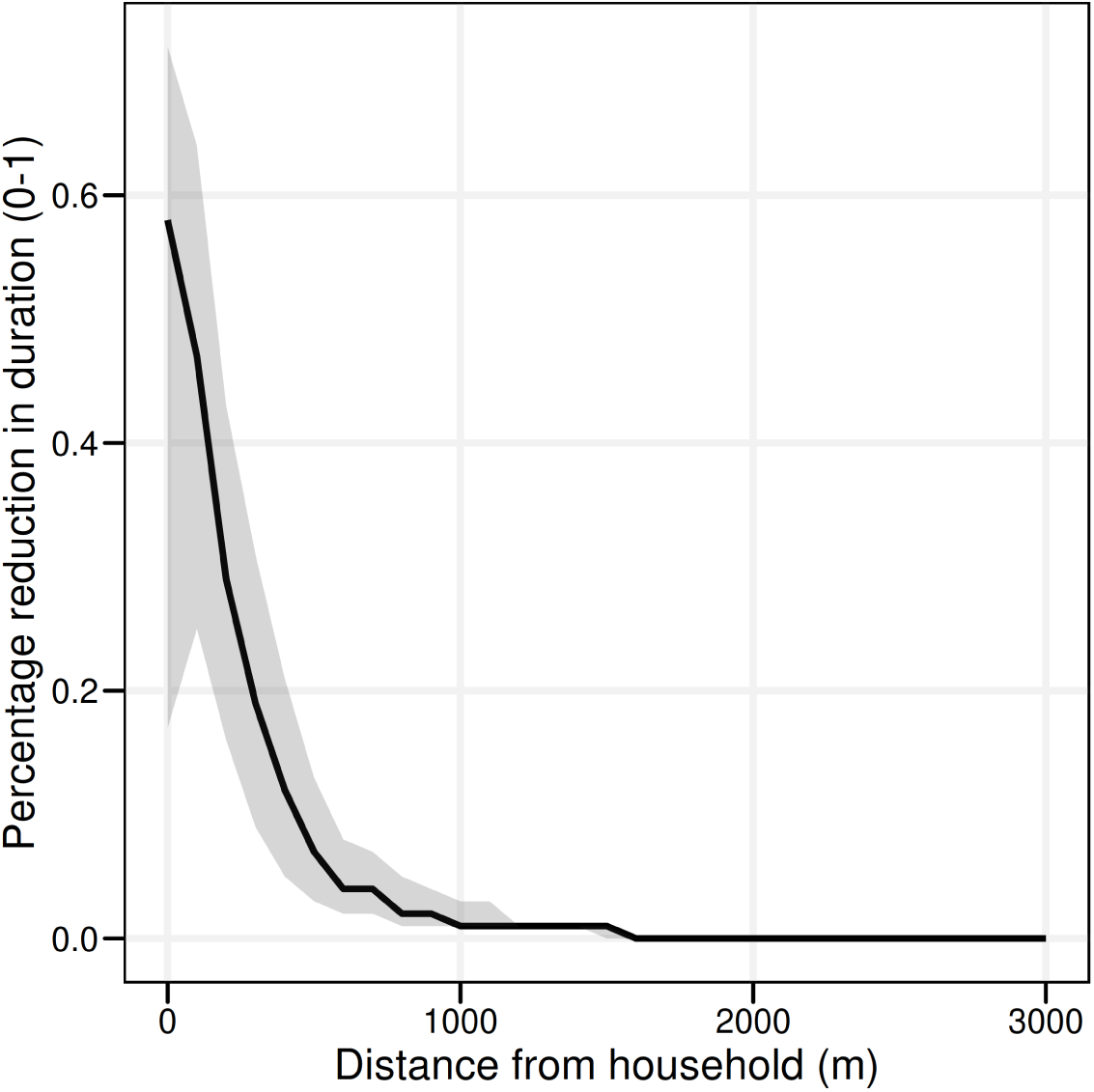
Crowding out of open water site usage duration over household distance. Estimated percentage reduction in the duration of open water site usage over household distance. This was estimated by comparing durations of open water site usage, estimated in the presence of taps/boreholes, to a counterfactual scenario where no taps/borehole were present, as shown in Fig. 7. Shaded grey areas represent 95% credible intervals.

**Fig. S7:**
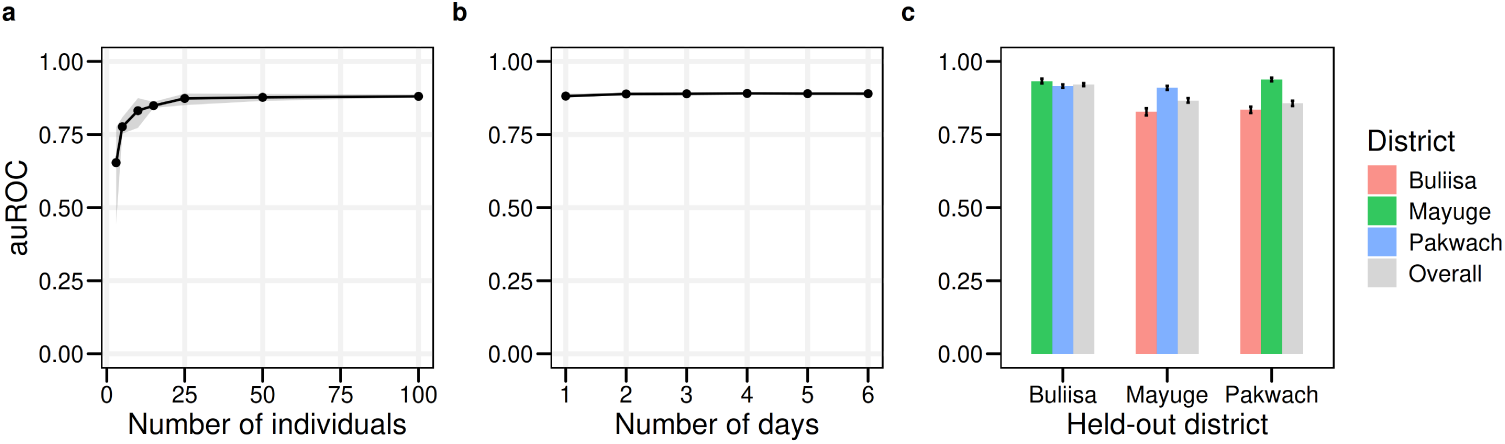
Minimum data needs for estimating the global spatial decay of water site usage. **a** auROC by number of individuals used to estimate the spatial decay. **b** auROC by number of GPS logger days used to estimate the spatial decay. **c** Evaluating performance on held-out districts. For (**a–b**) estimation was performed by randomly sampling from the training data three times and evaluating the auROC on held-out test data. For (**c**), models were estimated on the full data from one district and performance was evaluated separately for each of the held-out districts and for both held-out districts jointly (overall columns). The *x* -axis label in **(c)** indicates the held-out district. Shaded grey areas and vertical bars represent 95% credible intervals.

**Fig. S8:**
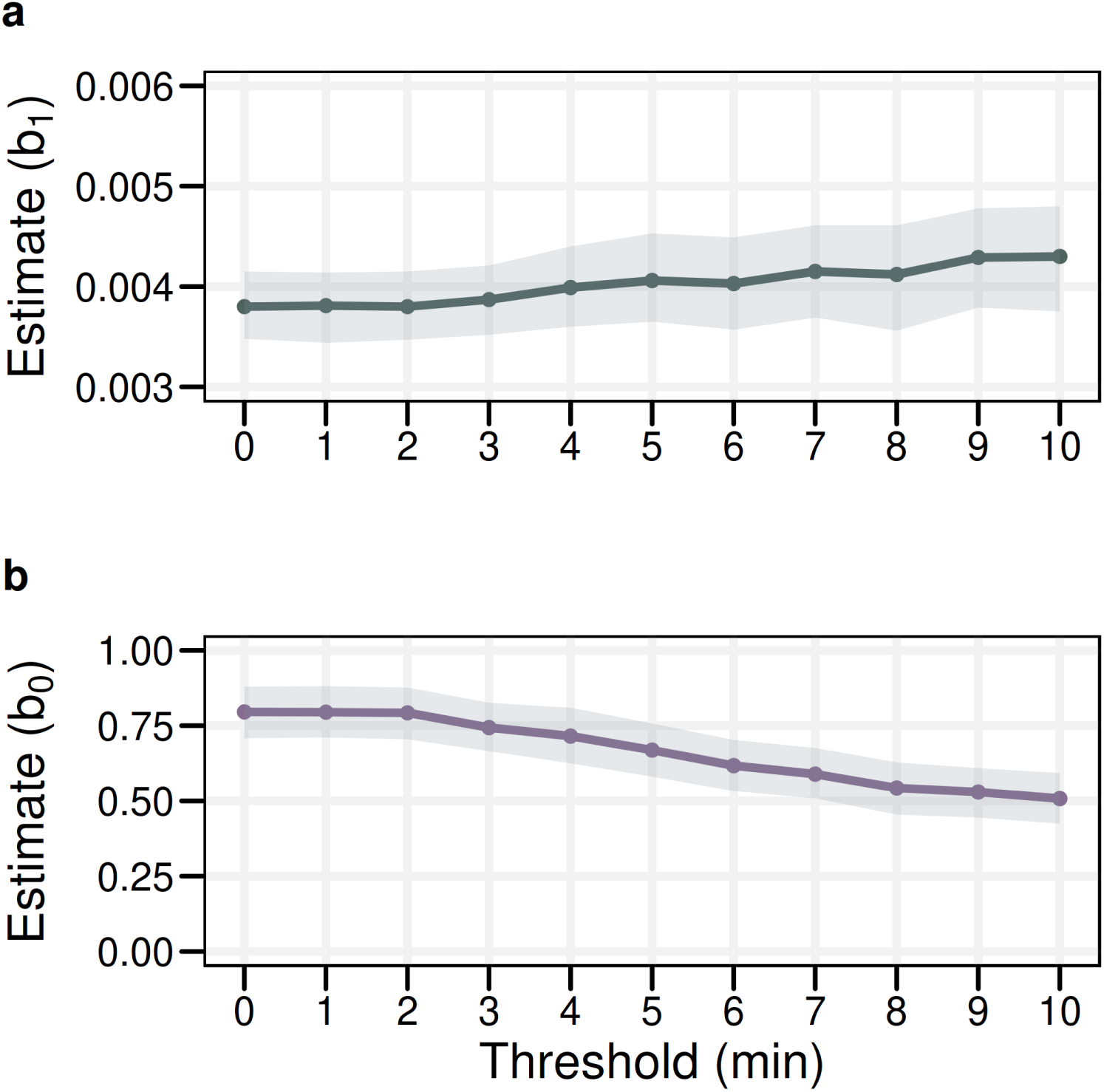
The impact of removing short-duration water site and tap/borehole usage events. The threshold value on the *x* -axis indicates the cut-off value below which events were removed (with a cut-off of zero denoting the dataset used for the main models where no events were removed). Shaded grey areas represent 95% credible intervals.

**Fig. S9:**
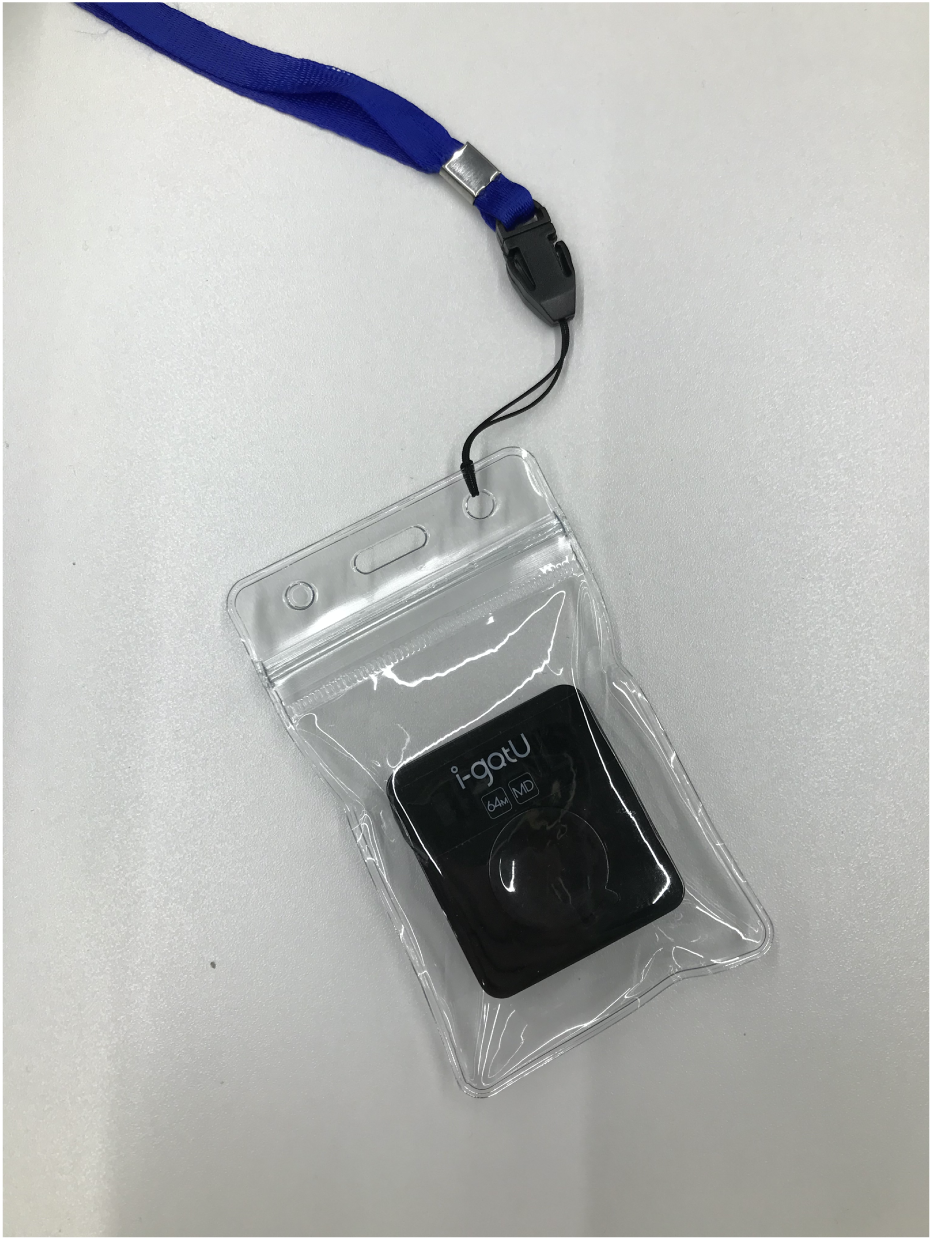
GPS logger in waterproof pouch and with attached lanyard.

**Fig. S10:**
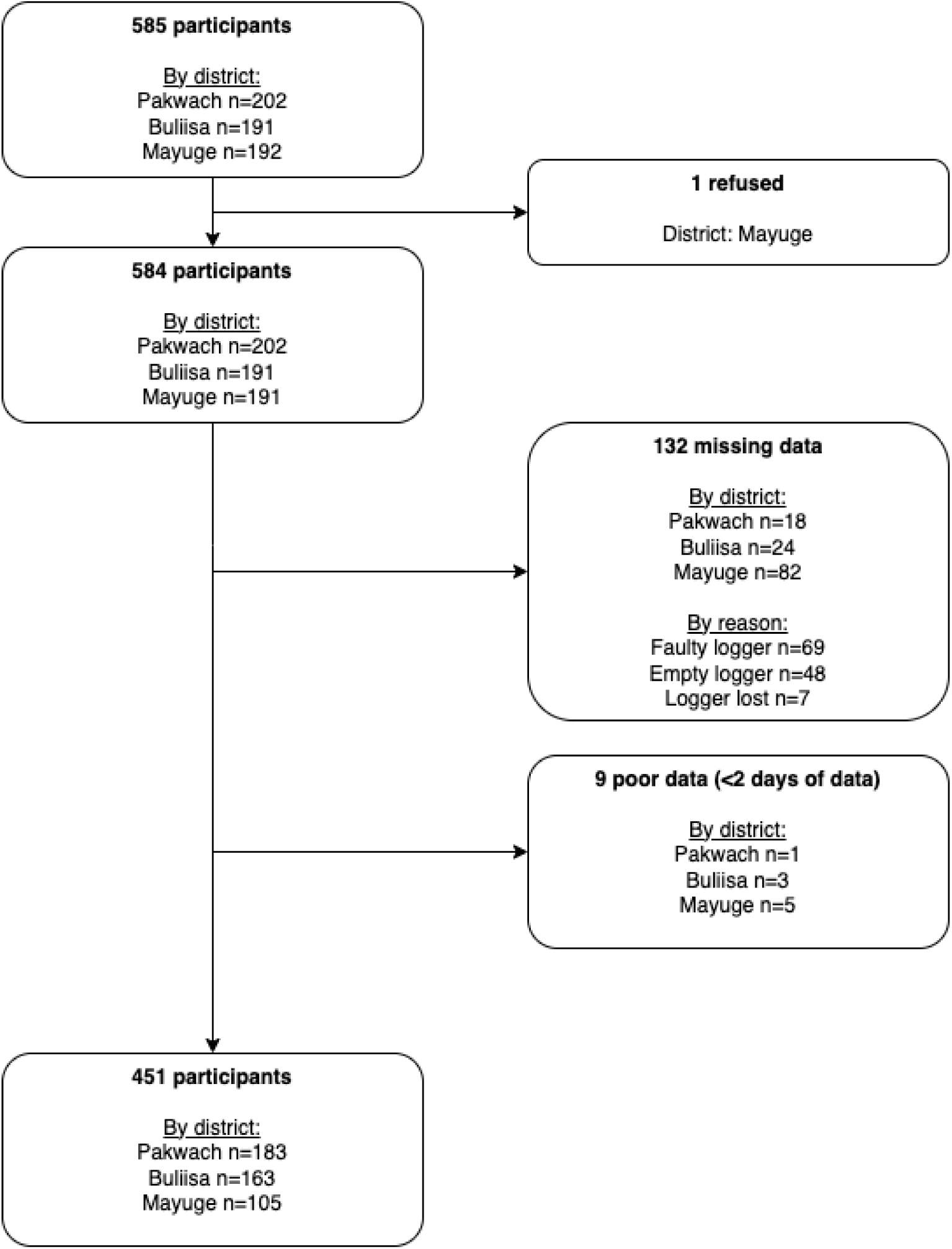
Participant flowchart. Loggers were classified as being faulty when no data could be retrieved due to complete device failure. Loggers were classified as being empty when loggers could still be pulled using a computer but contained no data.

**Fig. S11:**
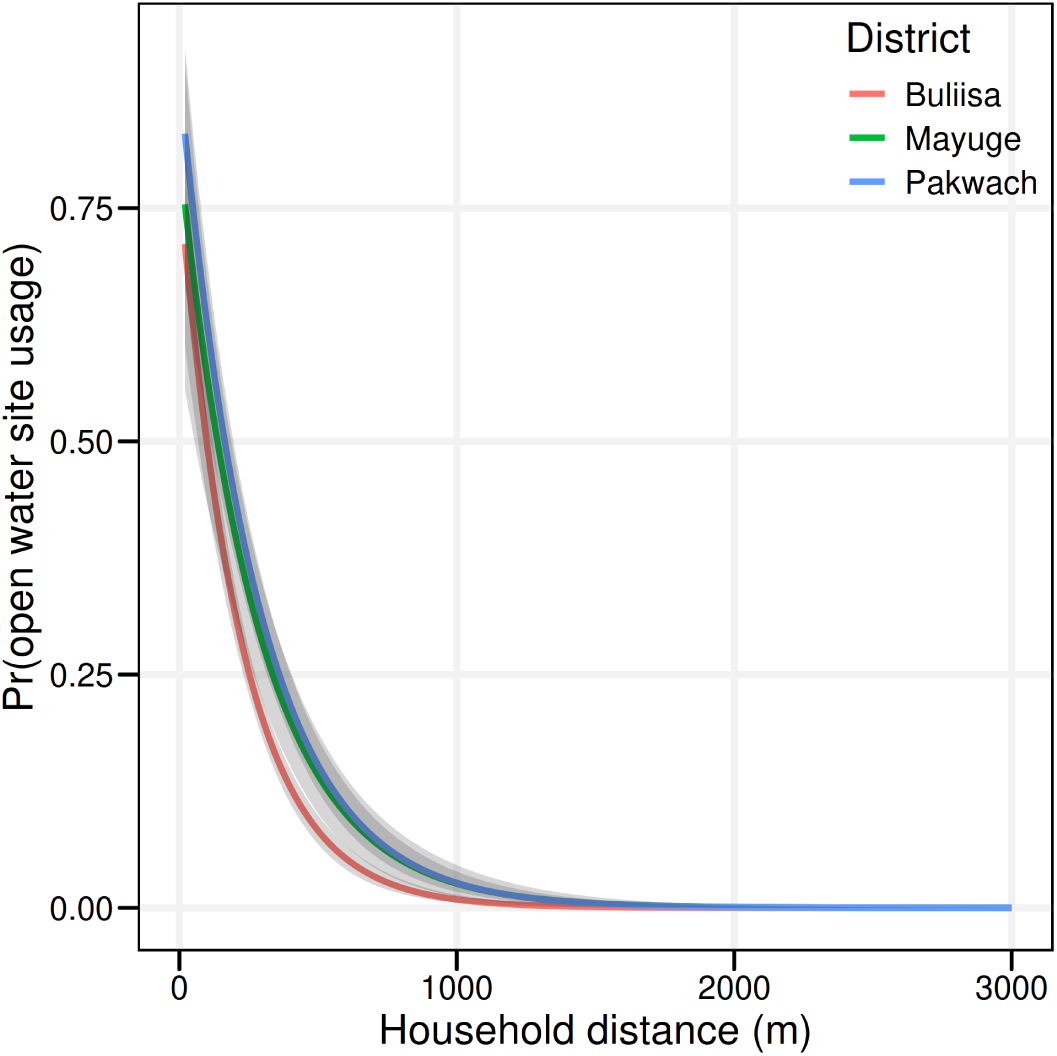
Spatial decay models of open water site usage incorporating a district-specific intercepts and decay functions. Shaded grey areas represent 95% credible intervals.

## Appendix C Supplementary tables

**Table S1:** Summary of key participant characteristics.

| Variable | Open water site usage |  |  | Tap/borehole usage |  |  |
| --- | --- | --- | --- | --- | --- | --- |
|  | No (n= 160) | Yes (n= 292) | P-value | No (n= 302) | Yes (n= 150) | P-value |
| Gender = male (%) | 83 (51.9) | 143 (49.0) | 0.62 | 150 (49.7) | 76 (50.7) | 0.92 |
| Age ≥ 15 years (%) | 86 (53.8) | 168 (57.5) | 0.50 | 171 (56.6) | 83 (55.3) | 0.87 |
| Occupation (%) |  |  | <0.01 |  |  | 0.60 |
| none | 95 (59.4) | 164 (56.2) |  | 176 (58.3) | 83 (55.3) |  |
| fishing | 7 (4.4) | 34 (11.6) |  | 24 (7.9) | 17 (11.3) |  |
| fishmongering | 2 (1.2) | 20 (6.8) |  | 15 (5.0) | 7 (4.7) |  |
| farming | 46 (28.7) | 47 (16.1) |  | 65 (21.5) | 28 (18.7) |  |
| other | 10 (6.2) | 27 (9.2) |  | 22 (7.3) | 15 (10.0) |  |
| Household drinking water source = improved (%) | 43 (26.9) | 183 (62.7) | <0.01 | 125 (41.4) | 101 (67.3) | <0.01 |
| Distance to the closest site (m) (median [IQR]) | 385.2 [248.3, 637.7] | 91.6 [58.9, 249.6] | <0.01 | 312.8 ± 292.8 | 209.6 ± 200.6 | <0.01 |
| Distance to the closest tap/borehole (m) (median [IQR]) | 396.2 [135.2, 715.2] | 179.4 [82.6, 404.9] | <0.01 | 515.7 ± 429.2 | 179.3 ± 270.7 | <0.01 |
| Radius of gyration (m) (median [IQR]) | 361.7 [170.9, 1180.5] | 351.5 [128.6, 1071.5] | 0.33 | 369.2 [152.6, 1092.6] | 314.8 [134.7, 1065.3] | 0.55 |

*Notes:* For binary variables, number and proportions [N (%)] are shown and Chi-squared tests were used to test for group differences. For all normally distributed continuous variables, mean and standard deviation (mean ± sd) are shown and two sample *t*-tests were performed to test for group differences. For all non-normally distributed variables, median and interquartile range (median [IQR]) are shown and Kruskal–Wallis Rank Sum Tests were performed to test for group differences.

**Table S2:** Logistic regression models predicting open water site usage.

| Type of model | Predictors | Sensitivity | Specificity | auROC |
| --- | --- | --- | --- | --- |
| Distance | Distance to water sites | 0.635 | 0.932 | 0.877 |
| Distance | Distance to water sites <430m (y/n) | 0.769 | 0.770 | 0.770 |
| Distance | Closest water site (y/n) | 0.462 | 0.936 | 0.699 |
| Socio-demographic proxies | Gender, age (<15, ≥ 15 years), occupation category, improved household drinking water source (y/n) | 0.500 | 0.682 | 0.594 |
| Behavioural proxies | Self-reported water contact (y/n), activity match (y/n), mobility tercile | 0.558 | 0.559 | 0.550 |

*Notes:* Bayesian logistic regression models using distance, socio-demographic proxies or behavioural proxies (see Methods) to predict open water site usage. Models were trained on 85% of the data and performance was evaluated on 15% of held-out data.

**Table S3:**
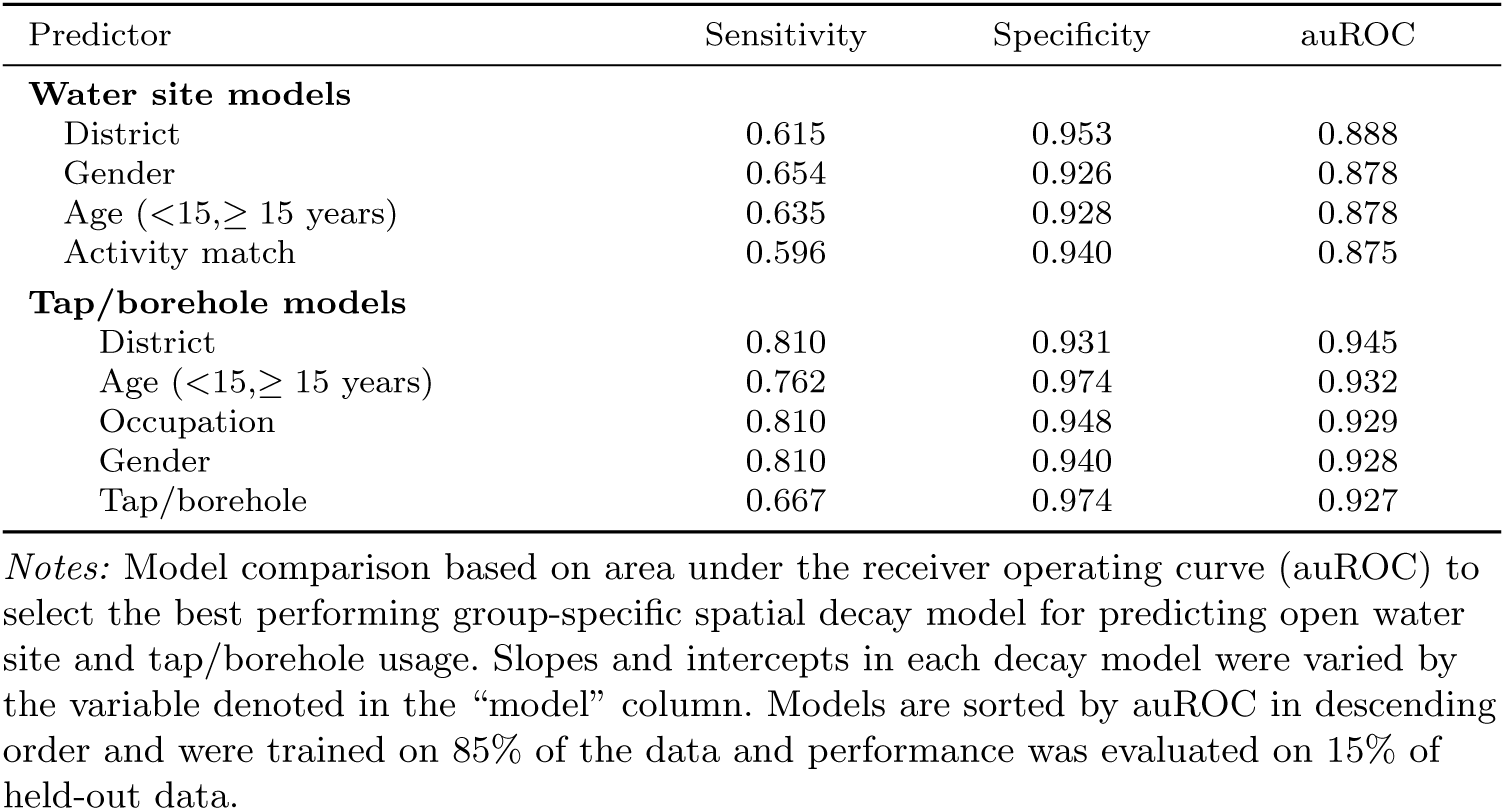
Model selection for spatial decay models.

**Table S4:**
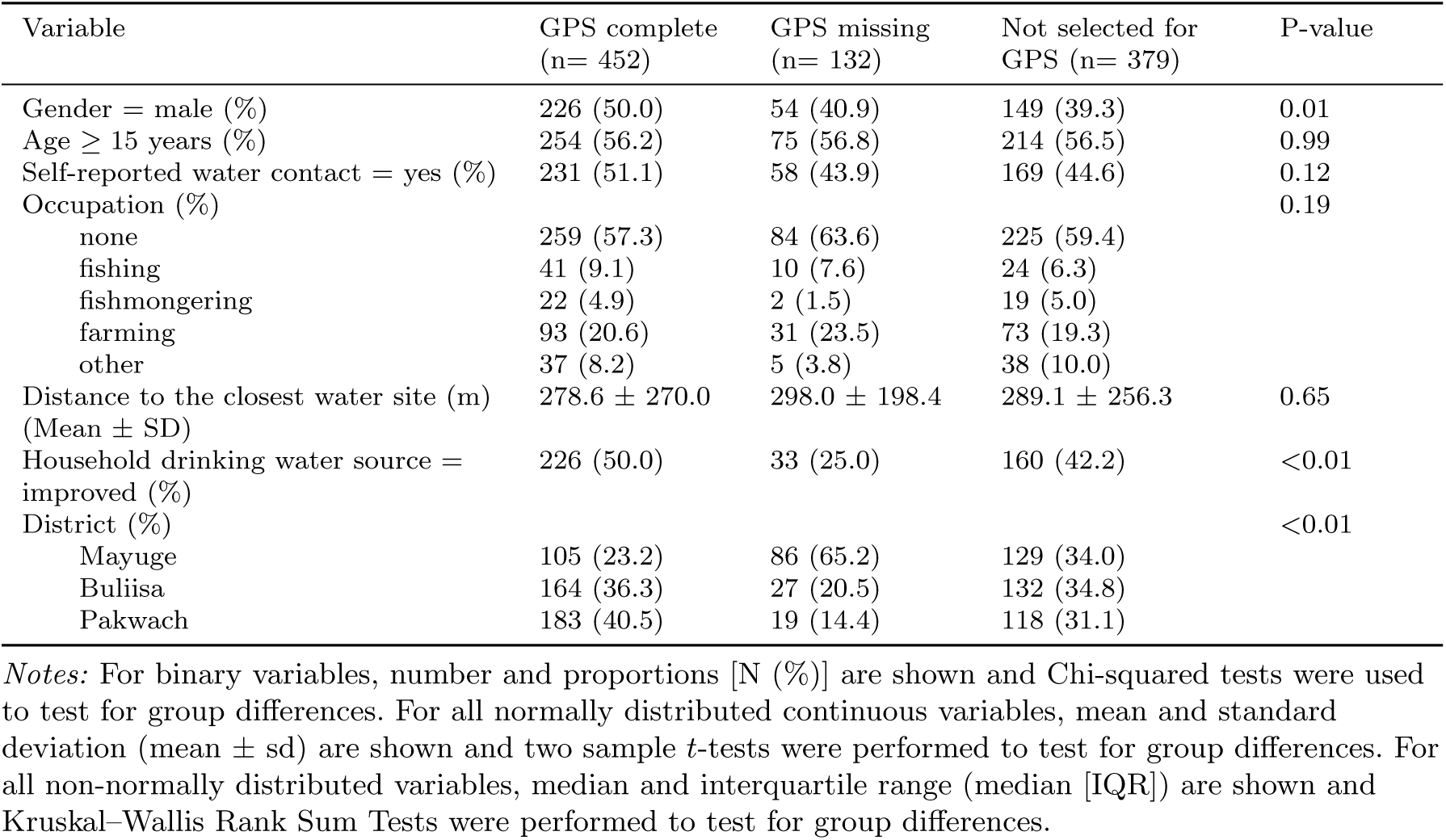
Comparison of participants with sufficient GPS data versus those with missing data and the wider SchistoTrack study population.

**Table S5:**
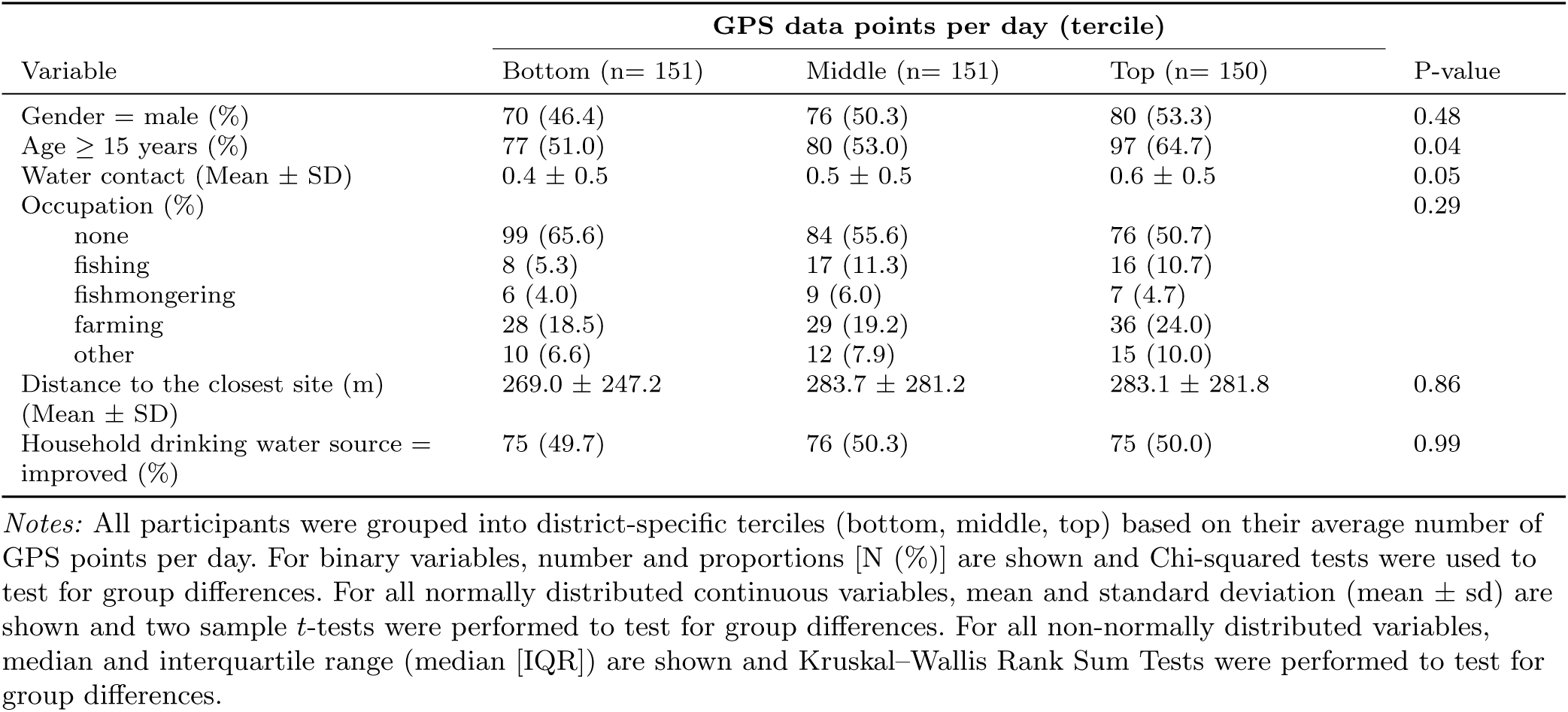
Comparison of participants by number of recorded GPS points, grouped into terciles.

**Table S6:**
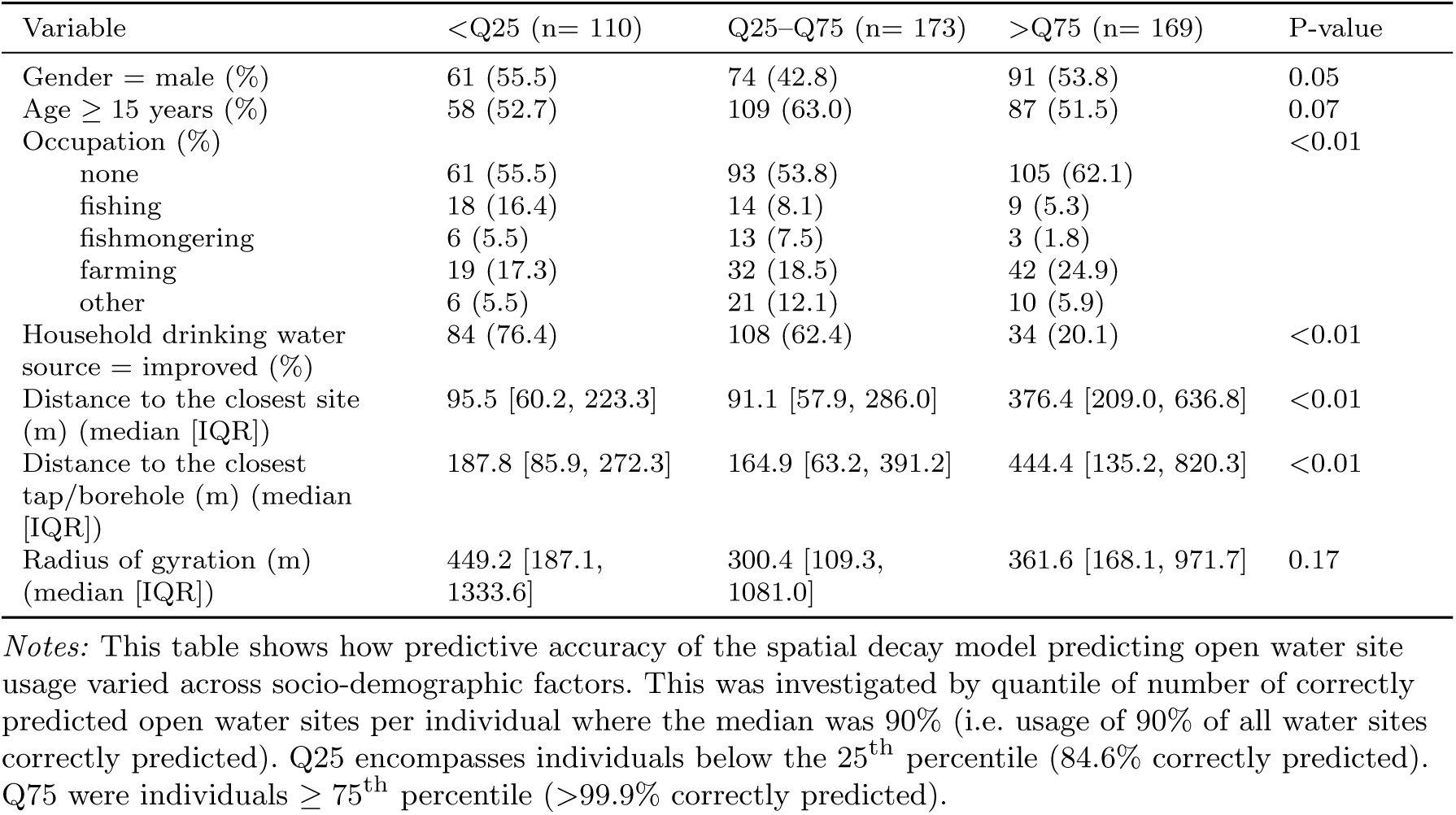
Differences in predictive performance across socio-demographic factors.

| Variable | <Q25 (n= 110) | Q25-Q75 (n= 173) | >Q75 (n= 169) | P-value |
| --- | --- | --- | --- | --- |
| Gender = male (%) | 61 (55.5) | 74 (42.8) | 91 (53.8) | 0.05 |
| Age ≥ 15 years (%) | 58 (52.7) | 109 (63.0) | 87 (51.5) | 0.07 |
| Occupation (%) |  |  |  | <0.01 |
| none | 61 (55.5) | 93 (53.8) | 105 (62.1) |  |
| fishing | 18 (16.4) | 14 (8.1) | 9 (5.3) |  |
| fishmongering | 6 (5.5) | 13 (7.5) | 3 (1.8) |  |
| farming | 19 (17.3) | 32 (18.5) | 42 (24.9) |  |
| other | 6 (5.5) | 21 (12.1) | 10 (5.9) |  |
| Household drinking water source = improved (%) | 84 (76.4) | 108 (62.4) | 34 (20.1) | <0.01 |
| Distance to the closest site (m) (median [IQR]) | 95.5 [60.2, 223.3] | 91.1 [57.9, 286.0] | 376.4 [209.0, 636.8] | <0.01 |
| Distance to the closest tap/borehole (m) (median [IQR]) | 187.8 [85.9, 272.3] | 164.9 [63.2, 391.2] | 444.4 [135.2, 820.3] | <0.01 |
| Radius of gyration (m) (median [IQR]) | 449.2 [187.1, 1333.6] | 300.4 [109.3, 1081.0] | 361.6 [168.1, 971.7] | 0.17 |

*Notes:* This table shows how predictive accuracy of the spatial decay model predicting open water site usage varied across socio-demographic factors. This was investigated by quantile of number of correctly predicted open water sites per individual where the median was 90% (i.e. usage of 90% of all water sites correctly predicted). Q25 encompasses individuals below the 25<sup>th</sup> percentile (84.6% correctly predicted). Q75 were individuals ≥ 75<sup>th</sup> percentile ( ≥ 99.9% correctly predicted).

## References

[1] Ferretti L, Wymant C, Petrie J, Tsallis D, Kendall M, Ledda A, et al. Digital measurement of SARS-CoV-2 transmission risk from 7 million contacts. Nature. 2023 Dec;626(7997):145–150. 10.1038/s41586-023-06952-2.

[2] Staedke SG, Nottingham EW, Cox J, Kamya MR, Rosenthal PJ, Dorsey G. Short report: Proximity to mosquito breeding sites as a risk factor for clinical malaria episodes in an urban cohort of Ugandan children. The American Journal of Tropical Medicine and Hygiene. 2003 Sep;69(3):244–246. 10.4269/ajtmh.2003.69.244.

[3] Arnold BF, Kanyi H, Njenga SM, Rawago FO, Priest JW, Secor WE, et al. Fine-scale heterogeneity in *Schistosoma mansoni* force of infection measured through antibody response. Proceedings of the National Academy of Sciences. 2020 Aug;117(37):23174–23181. 10.1073/pnas.2008951117.

[4] Wesolowski A, Eagle N, Tatem AJ, Smith DL, Noor AM, Snow RW, et al. Quantifying the Impact of Human Mobility on Malaria. Science. 2012 Oct;338(6104):267–270. 10.1126/science.1223467.

[5] Stoddard ST, Forshey BM, Morrison AC, Paz-Soldan VA, Vazquez-Prokopec GM, Astete H, et al. House-to-house human movement drives dengue virus transmission. Proceedings of the National Academy of Sciences. 2012 Dec;110(3):994–999. 10.1073/pnas.1213349110.

[6] Buckee C, Noor A, Sattenspiel L. Thinking clearly about social aspects of infectious disease transmission. Nature. 2021 Jun;595(7866):205–213. 10.1038/s41586-021-03694-x.

[7] Jia JS, Lu X, Yuan Y, Xu G, Jia J, Christakis NA. Population flow drives spatio-temporal distribution of COVID-19 in China. Nature. 2020 Apr;582(7812):389–394. 10.1038/s41586-020-2284-y.

[8] Kostandova N, Schluth C, Arambepola R, Atuhaire F, Bérubé S, Chin T, et al. A systematic review of using population-level human mobility data to understand SARS-CoV-2 transmission. Nature Communications. 2024 Dec;15(1). 10.1038/s41467-024-54895-7.

[9] Wolfe ND, Dunavan CP, Diamond J. Origins of major human infectious diseases. Nature. 2007 May;447(7142):279–283. 10.1038/nature05775.

[10] Findlater A, Bogoch II. Human Mobility and the Global Spread of Infectious Diseases: A Focus on Air Travel. Trends in Parasitology. 2018 Sep;34(9):772–783. 10.1016/j.pt.2018.07.004.

[11] Colizza V, Barrat A, Barthélemy M, Vespignani A. The role of the airline transportation network in the prediction and predictability of global epidemics. Proceedings of the National Academy of Sciences. 2006 Feb;103(7):2015–2020. 10.1073/pnas.0510525103.

[12] Brockmann D, Helbing D. The Hidden Geometry of Complex, Network-Driven Contagion Phenomena. Science. 2013 Dec;342(6164):1337–1342. 10.1126/science.1245200.

[13] World Health Organization Control of Neglected Tropical Diseases Team.: Schistosomiasis and Soil-Transmitted Helminthiases: Progress Report, 2023 [Weekly Epidemiological Record]. https://www.who.int/publications/i/item/WHO-WER9948-707-717.

[14] Grimes JET, Croll D, Harrison WE, Utzinger J, Freeman MC, Templeton MR. The Relationship between Water, Sanitation and Schistosomiasis: A Systematic Review and Meta-analysis. PLoS Neglected Tropical Diseases. 2014 12;8(12):e3296. 10.1371/journal.pntd.0003296.

[15] Reitzug F, Ledien J, Chami GF. Associations of water contact frequency, duration, and activities with schistosome infection risk: A systematic review and meta-analysis. PLoS Neglected Tropical Diseases. 2023 Jun;17(6):e0011377. 10.1371/journal.pntd.0011377.

[16] Woolhouse MEJ, Dye C, Etard JF, Smith T, Charlwood JD, Garnett GP, et al. Heterogeneities in the transmission of infectious agents: Implications for the design of control programs. Proceedings of the National Academy of Sciences. 1997 1;94(1):338–342. 10.1073/pnas.94.1.338.

[17] Woolhouse M, Etard JF, Dietz K, Ndhlovu P, Chandiwana S. Heterogeneities in schistosome transmission dynamics and control. Parasitology. 1998;117(5):475–482.

[18] Kraemer MUG, Sadilek A, Zhang Q, Marchal NA, Tuli G, Cohn EL, et al. Mapping global variation in human mobility. Nature Human Behaviour. 2020 May;4(8):800–810. 10.1038/s41562-020-0875-0.

[19] Mari L, Gatto M, Ciddio M, Dia ED, Sokolow SH, De Leo GA, et al. Bigdata-driven modeling unveils country-wide drivers of endemic schistosomiasis. Scientific Reports. 2017 Mar;7(1). 10.1038/s41598-017-00493-1.

[20] Ciddio M, Mari L, Sokolow SH, De Leo GA, Casagrandi R, Gatto M. The spatial spread of schistosomiasis: A multidimensional network model applied to Saint-Louis region, Senegal. Advances in Water Resources. 2017 Oct;108:406–415. 10.1016/j.advwatres.2016.10.012.

[21] Hardwick RJ, Vegvari C, Collyer B, Truscott JE, Anderson RM. Spatial scales in human movement between reservoirs of infection. Journal of Theoretical Biology. 2021 Sep;524:110726. 10.1016/j.jtbi.2021.110726.

[22] Perez-Saez J, Mari L, Bertuzzo E, Casagrandi R, Sokolow SH, De Leo GA, et al. A Theoretical Analysis of the Geography of Schistosomiasis in Burkina Faso Highlights the Roles of Human Mobility and Water Resources Development in Disease Transmission. PLOS Neglected Tropical Diseases. 2015 Oct;9(10):e0004127. 10.1371/journal.pntd.0004127.

[23] Kloos H, Correa-Oliveira R, Reis DCd, Rodrigues EW, Monteiro LAS, Gazzinelli A. The role of population movement in the epidemiology and control of schistosomiasis in Brazil: a preliminary typology of population movement. Memórias do Instituto Oswaldo Cruz. 2010 Jul;105(4):578–586. 10.1590/s0074-02762010000400038.

[24] Reitzug F, Kabatereine NB, Byaruhanga AM, Besigye F, Nabatte B, Chami GF. Current water contact and *Schistosoma mansoni* infection have distinct determinants: a data-driven population-based study in rural Uganda. Nature Communications. 2024 Nov;15(1). 10.1038/s41467-024-53519-4.

[25] Lamberti O, Kabatereine NB, Tukahebwa EM, Chami GF. *Schistosoma mansoni* infection risk for school-aged children clusters within households and is modified by distance to freshwater bodies. PLoS ONE. 2021 Nov;16(11):e0258915. 10.1371/journal.pone.0258915.

[26] Kloos H, Fulford AJC, Butterworth AE, Sturrock RF, Ouma JH, Kariuki HC, et al. Spatial patterns of human water contact and Schistosoma mansoni transmission and infection in four rural areas in Machakos District, Kenya. Social Science & Medicine. 1997 Apr;44(7):949–968. 10.1016/s0277-9536(96)00218-3.

[27] World Health Organization. WHO guideline on control and elimination of human schistosomiasis. Geneva: World Health Organization; 2022. Section: xxi, 108 p. Available from: https://apps.who.int/iris/handle/10665/351856.

[28] World Health Organization.: Field use of molluscicides in schistosomiasis control programmes: an operational manual for programme managers. Geneva: World Health Organization. Available from: https://apps.who.int/iris/handle/10665/255644.

[29] Nuffield Department of Population Health, University of Oxford.: SchistoTrack: a prospective multimorbidity cohort. 2024-04-16. https://www.bdi.ox.ac.uk/research/schistotrack.

[30] Iacovidou MA, Byaruhanga AM, Besigye F, Nabatte B, Kabatereine NB, Chami GF. Ecological niche stability ofBiomphalariaintermediate hosts for *Schistosoma mansoni* under extreme flooding and seasonal change. medRxiv. 2025 Jan;10.1101/2025.01.07.631649.

[31] Zipf GK. The P 1 P 2/D hypothesis: on the intercity movement of persons. American sociological review. 1946;11(6):677–686.

[32] Simini F, González MC, Maritan A, Barabási AL. A universal model for mobility and migration patterns. Nature. 2012 Feb;484(7392):96–100. 10.1038/nature10856.

[33] Eyre MT, Stanton MC, Macklin G, Bartoníček Z, O’Halloran L, Eloundou Ombede DR, et al. Piloting an integrated approach for estimation of environmental risk of *Schistosoma haematobium* infections in pre-school-aged children and their mothers at Barombi Kotto, Cameroon. Acta Tropica. 2020 Dec;212:105646. 10.1016/j.actatropica.2020.105646.

[34] Ruiz Cuenca P, Souza FN, Coutinho do Nascimento R, Goncalves da Silva A, Eyre MT, Santana JO, et al. Using step selection functions to analyse human mobility using telemetry data in infectious disease epidemiology: a case study of leptospirosis. medRxiv. 2025 May;10.1101/2025.04.28.25326582.

[35] González MC, Hidalgo CA, Barabási AL. Understanding individual human mobility patterns. Nature. 2008 Jun;453(7196):779–782. 10.1038/nature06958.

[36] Pappalardo L, Simini F, Rinzivillo S, Pedreschi D, Giannotti F, Barabási AL. Returners and explorers dichotomy in human mobility. Nature Communications. 2015 Sep;6(1). 10.1038/ncomms9166.

[37] Wesolowski A, O’Meara WP, Eagle N, Tatem AJ, Buckee CO. Evaluating Spatial Interaction Models for Regional Mobility in Sub-Saharan Africa. PLOS Computational Biology. 2015 Jul;11(7):e1004267. 10.1371/journal.pcbi.1004267.

[38] Meredith HR, Giles JR, Perez-Saez J, Mande T, Rinaldo A, Mutembo S, et al. Characterizing human mobility patterns in rural settings of sub-Saharan Africa. eLife. 2021 Sep;10. 10.7554/elife.68441.

[39] Seto EYW, Sousa-Figueiredo JC, Betson M, Byalero C, Kabatereine NB, Stothard JR. Patterns of intestinal schistosomiasis among mothers and young children from Lake Albert, Uganda: water contact and social networks inferred from wearable global positioning system dataloggers. Geospatial health. 2012 11;7(1):1. 10.4081/gh.2012.99.

[40] Cairncross S. Trachoma and water. Community eye health. 1999;12(32):58.

[41] Sangare M, Diabate AF, Coulibaly YI, Tanapo D, Thera SO, Dolo H, et al. Understanding the barriers and facilitators related to never treatment during mass drug administration among mobile and migrant populations in Mali: a qualitative exploratory study. BMJ Global Health. 2024 Oct;9(10):e015671. 10.1136/bmjgh-2024-015671.

[42] Adams MW, Sutherland EG, Eckert EL, Saalim K, Reithinger R. Leaving no one behind: targeting mobile and migrant populations with health interventions for disease elimination—a descriptive systematic review. BMC Medicine. 2022 May;20(1). 10.1186/s12916-022-02365-6.

[43] Parker M, Allen T, Pearson G, Peach N, Flynn R, Rees N. Border parasites: schistosomiasis control among Uganda’s fisherfolk. Journal of Eastern African Studies. 2012 Feb;6(1):98–123. 10.1080/17531055.2012.664706.

[44] Graham M, Ayabina D, Lucas TC, Collyer BS, Medley GF, Hollingsworth TD, et al. SCHISTOX: An individual based model for the epidemiology and control of schistosomiasis. Infectious Disease Modelling. 2021;6:438–447. 10.1016/j.idm.2021.01.010.

[45] Anderson RM, Turner HC, Farrell SH, Truscott JE. In: Studies of the Transmission Dynamics, Mathematical Model Development and the Control of Schistosome Parasites by Mass Drug Administration in Human Communities. Elsevier; 2016. p. 199–246. Available from: 10.1016/bs.apar.2016.06.003.

[46] Knopp S, Person B, Ame SM, Ali SM, Hattendorf J, Juma S, et al. Evaluation of integrated interventions layered on mass drug administration for urogenital schistosomiasis elimination: a cluster-randomised trial. The Lancet Global Health. 2019 Aug;7(8):e1118–e1129. 10.1016/s2214-109x(19)30189-5.

[47] World Health Organization. WASH and Health Working Together: A ‘how-to’ Guide for Neglected Tropical Disease Programmes. World Health Organization; 2023.

[48] Google, Microsoft.: Google-Microsoft Open Buildings - combined by VIDA. 2024-12-30. https://beta.source.coop/repositories/vida/google-microsoft-open-buildings.

[49] Tatem AJ. WorldPop, open data for spatial demography. Scientific Data. 2017 Jan;4(1). 10.1038/sdata.2017.4.

[50] Sow S, de Vlas SJ, Stelma F, Vereecken K, Gryseels B, Polman K. The contribution of water contact behavior to the high *Schistosoma mansoni* Infection rates observed in the Senegal River Basin. BMC Infectious Diseases. 2011 12;11(1):198. 10.1186/1471-2334-11-198.

[51] Sedgwick P. The Hawthorne effect. BMJ. 2011 Jan; 344(jan04 2):d8262–d8262. 10.1136/bmj.d8262.

[52] Fleming CH, Calabrese JM.: ctmm: Continuous-Time Movement Modeling. Available from: https://github.com/ctmm-initiative/ctmm,https://groups.google.com/g/ctmm-user.

[53] Morris G, Conner LM. Assessment of accuracy, fix success rate, and use of estimated horizontal position error (EHPE) to filter inaccurate data collected by a common commercially available GPS logger. PLoS ONE. 2017;12(11):e0189020. 10.1371/journal.pone.0189020.

[54] Berger-Tal O, Bar-David S. Recursive movement patterns: review and synthesis across species. Ecosphere. 2015 Sep;6(9):1–12. 10.1890/ es15-00106.1.

[55] Seidel DP, Dougherty E, Carlson C, Getz WM. Ecological metrics and methods for GPS movement data. International Journal of Geographical Information Science. 2018 Jul;32(11):2272–2293. 10.1080/13658816.2018.1498097.

[56] Bracis C.: recurse: Computes Revisitation Metrics for Trajectory Data. https://CRAN.R-project.org: Comprehensive R Archive Network (CRAN). Version 1.4.0. Available from: https://CRAN.R-project.org/package=recurse.

[57] Hahsler M, Piekenbrock M, Doran D. dbscan: Fast Density-Based Clustering with R. Journal of Statistical Software. 2019;91(1). 10.18637/jss.v091.i01.

[58] Kwiringira J, Atekyereza P, Niwagaba C, Günther I. Gender variations in access, choice to use and cleaning of shared latrines; experiences from Kampala Slums, Uganda. BMC Public Health. 2014 Nov;14(1). 10.1186/1471-2458-14-1180.

[59] Tessema RA. Assessment of the implementation of community-led total sanitation, hygiene, and associated factors in Diretiyara district, Eastern Ethiopia. PLOS ONE. 2017 Apr;12(4):e0175233. 10.1371/journal.pone.0175233.

[60] Webster BL, Diaw OT, Seye MM, Faye DS, Stothard JR, Sousa-Figueiredo JC, et al. Praziquantel treatment of school children from single and mixed infection foci of intestinal and urogenital schistosomiasis along the Senegal River Basin: monitoring treatment success and re-infection patterns. Acta Tropica. 2013 Nov;128(2):292–302. 10.1016/j.actatropica.2012.09.010.

[61] Tchuem Tchuenté LA, Momo SC, Stothard JR, Rollinson D. Efficacy of praziquantel and reinfection patterns in single and mixed infection foci for intestinal and urogenital schistosomiasis in Cameroon. Acta Tropica. 2013 Nov;128(2):275–283. 10.1016/j.actatropica.2013.06.007.

[62] Buuren Sv, Groothuis-Oudshoorn K. mice: Multivariate Imputation by Chained Equations inR. Journal of Statistical Software. 2011;45(3). 10.18637/jss.v045.i03.

[63] World Resources Institute.: Waterbodies in Uganda. [accessed 16 Apr 2024]. https://datasets.wri.org/dataset/waterbodies-in-uganda.

[64] Vehtari A, Gelman A, Gabry J. Practical Bayesian model evaluation using leave-one-out cross-validation and WAIC. Statistics and Computing. 2016 Aug;27(5):1413–1432. 10.1007/s11222-016-9696-4.

[65] Stouffer SA. Intervening Opportunities: A Theory Relating Mobility and Distance. American Sociological Review. 1940 Dec;5(6):845. 10.2307/2084520.

[66] Bürkner PC. brms: An R Package for Bayesian Multilevel Models Using Stan. Journal of Statistical Software. 2017;80(1). 10.18637/jss.v080.i01.

[67] Hoffman MD, Gelman A, et al. The No-U-Turn sampler: adaptively setting path lengths in Hamiltonian Monte Carlo. Journal of Machine Learning Research. 2014;15(1):1593–1623.

[68] Thiele C, Hirschfeld G. cutpointr: Improved Estimation and Validation of Optimal Cutpoints in R. Journal of Statistical Software. 2021;98(11):1–27. 10.18637/jss.v098.i11.

[69] Yang Y, Herrera C, Eagle N, González MC. Limits of Predictability in Commuting Flows in the Absence of Data for Calibration. Scientific Reports. 2014 Jul;4(1). 10.1038/srep05662.

[70] Masucci AP, Serras J, Johansson A, Batty M. Gravity versus radiation models: On the importance of scale and heterogeneity in commuting flows. Physical Review E. 2013 Aug;88(2). 10.1103/physreve.88.022812.

[71] Li W, Wang Q, Liu Y, Small ML, Gao J. A spatiotemporal decay model of human mobility when facing large-scale crises. Proceedings of the National Academy of Sciences. 2022 Aug;119(33). 10.1073/pnas.2203042119.

